# Proteomic signatures of the *APOE ε4* and *APOE ε2* genetic variants and Alzheimer’s disease

**DOI:** 10.1101/2025.08.04.25332945

**Authors:** Lina Lu, Alexa Pichet Binette, Ines Hristovska, Shorena Janelidze, Bart Smets, Irene Cumplido Mayoral, Aparna Vasanthakumar, Britney Milkovich, Rik Ossenkoppele, Varsha Krish, Farhad Imam, Sebastian Palmqvist, Jacob Vogel, Erik Stomrud, Oskar Hansson, Niklas Mattsson-Carlgren

## Abstract

The ε4 and ε2 alleles of the Apolipoprotein E (*APOE*) gene confer opposite genetic risks for Alzheimer’s disease (AD), but their underlying molecular mechanisms remain poorly characterized in humans. To resolve this, we systematically profiled *APOE*-associated proteomic alterations across five cohorts—including the Global Neurodegeneration Proteomics Consortium (GNPC), BioFINDER-2, the Alzheimer’s Disease Neuroimaging Initiative (ADNI), the Parkinson’s Progression Markers Initiative (PPMI), and UK Biobank (UKB)—using SomaLogic and OLINK platforms in plasma and cerebrospinal fluid (CSF) from over 10,000 individuals. Using GNPC (plasma SomaLogic, N=4,045), we mapped a comprehensive *APOE*-protein network and applied mediation modeling to classify genotype-related signals as upstream mediators, downstream consequences, or *APOE*-specific changes. We then leveraged CSF beta-amyloid (Aβ) biomarker data from BioFINDER-2 (plasma SomaLogic, N=1,421) to improve temporal resolution and isolate early, Aβ-independent proteomic programs. In the Aβ-individuals, *APOE4* was linked to cell cycle and chromatin remodeling, while *APOE2* was associated with mitochondrial regulation and DNA repair. Mediation analyses nominated proteins such as S100A13, TBCA, SPC25 for *APOE4*, and APOB, SNAP23 for *APOE2* as candidate upstream effectors, supported by CSF validation (ADNI, SomaLogic, N=666), brain transcriptomic co-expression, and AD GWAS colocalization. Longitudinal CSF data from PPMI confirmed the temporal stability of several *APOE*-associated proteins. Cross-platform comparisons (UKB plasma OLINK, N=4,820, and BF2 CSF OLINK, N=1,475) revealed matrix- and assay-specific heterogeneity, underscoring challenges in reproducibility. Together, our results delineate allele-specific, temporally structured proteomic signatures that precede AD pathology, offering insight into *APOE*-driven molecular pathways and potential therapeutic targets for early intervention.

## Main

The Apolipoprotein E (*APOE*) gene is the strongest genetic factor for sporadic Alzheimer’s disease (AD), with three main alleles: ε2, ε3, and ε4. Compared to ε3, the ε4 allele (*APOE4*) increases AD risk in a dose-dependent manner—approximately 2–3 fold in people with one ε4 allele and up to 12-fold in ε4 homozygotes^1,2^ and is linked to Aβ aggregation^3–5^. In contrast, *APOE2* carriers have a reduced AD risk^6–8^ and Aβ burden^9–11^, and delayed onset in high-penetrance AD mutations carriers^12^. *APOE* has been suggested to contribute to AD via lipid metabolism^13,14^, mitochondrial function^15,16^, Aβ regulation^17,18^, and neuroinflammation^19,20^, but most data on *APOE* comes from animal or cell experiments, with limited human evidence supporting specific molecular pathways linking *APOE* to AD. Developments in proteomics provide a new perspective for analyzing the molecular heterogeneity of AD, for example identifying CSF biomarkers in both asymptomatic and symptomatic AD^21^, and CSF biomarkers related to Aβ- and tau-related pathology^22^. Proteomic studies also suggested *APOE4*-dependent pathways^23^, but were generally limited by small sample sizes^24,25^, or reliance on single tissue sources or single proteomics platforms^23,26,27^, limiting generalizability. To reduce bias from individual methods or tissues, there is a need for integrative studies that combine multiple cohorts, matrices (e.g., CSF and blood), and analytical platforms. A systematic, fine-grained analysis of the *APOE*-related protein network—considering both *APOE2* and *APOE4*—may reveal distinct mechanistic pathways and inform targeted intervention strategies.

Therefore, we mapped *APOE*-related proteins across large cohorts and tissues using multi-level proteomics, revealing distinct *APOE4*- and *APOE2*-associated signatures linked to AD pathology. This provides a molecular basis for future stratified management and targeted therapies.

## Results

### The APOE-plasma protein signature and its role in AD: Discovery cohort

#### The GNPC plasma SomaLogic Cohort

In the Global Neurodegeneration Proteomics Consortium (GNPC) cohort (N=4,045, aged 20-90, median age: 75), *APOE4*-associated proteins were identified by comparing *APOE4* carriers (1,334 ε3ε4; 243 ε4ε4) to ε3ε3 individuals (N=1,969). To assess the protective effect of *APOE2* in AD, ε2 carriers (13 ε2ε2; 375 ε2ε3) were compared with ε3ε3 individuals. ε2ε4 carriers (N=111) were excluded from these comparisons due to potential confounding but included in analyses of protein level changes across all *APOE* genotypes (Supplement Table 1).

#### *APOE4* and AD protein signatures: variances and similarity

To delineate the distinct and overlapping proteomic landscapes of *APOE4* and AD (clinical diagnosis), we applied three regression models across 7,285 proteins. Model 1 identified 478 proteins associated with *APOE4* (unadjusted for AD diagnosis), e.g., upregulated SPC25 and LRRN1, and downregulated S100A13 and TBCA (Fig. 2a). Model 2 identified 4,014 proteins associated with AD (unadjusted for *APOE4*), with top hits e.g., ACHE, SPC25 and MMP8 (Fig. 2b). Model 3 included terms for both *APOE4* and AD to isolate independent effects (Supplement Fig. 1A, B). Of the *APOE4*-associated proteins, 253 (53%) remained significant after adjustment for AD diagnosis, including 78 not linked to AD—suggesting *APOE4*-specific pathways (Fig. 2c, red). Conversely, 3,783 (94%) AD-associated proteins remained significant after adjustment for *APOE4* status, with 3,453 specific to AD (Fig. 2c, blue). 98 proteins were independently associated with both *APOE4* and AD across models (Fig. 2c, black).

**Fig. 1:**
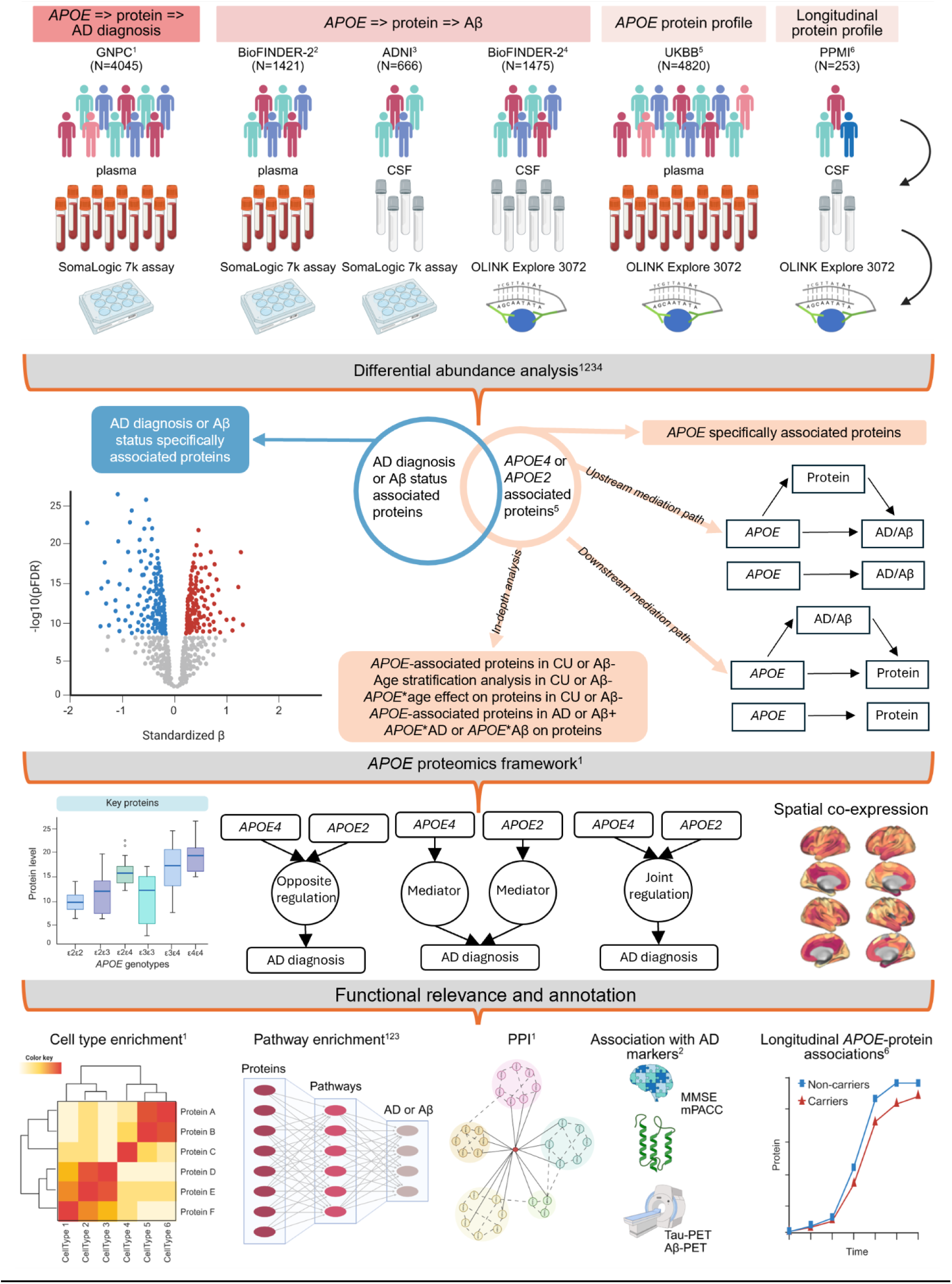
Study design. Overview of datasets: The Global Neurodegeneration Proteomics Consortium (GNPC) cohort (N=4,045, plasma, SomaLogic 7k) was used to identify proteins associated with *APOE4* or *APOE2* and clinical diagnosis (CU vs. AD dementia). To investigate *APOE*-related proteomic changes in relation to Aβ status (Aβ- vs. Aβ+), we utilized plasma data from BioFINDER-2 (SomaLogic 7k, a subset of GNPC), CSF data from Alzheimer’s Disease Neuroimaging Initiative (ADNI) (SomaLogic 7k), and CSF data from BioFINDER-2 (OLINK). The UK BioBank (UKB) (plasma, OLINK) was used exclusively to define *APOE*-driven protein signatures, as clinical diagnosis and Aβ status were unavailable in the dataset. Parkinson’s Progression Markers Initiative (PPMI) cohort (CSF, OLINK) is utilized to investigate the longitudinal *APOE*-protein associations. Differential abundance analysis identified *APOE* (*APOE4* or *APOE2*) associated proteins and AD diagnosis (or Aβ status) associated proteins, resulting in different groups of proteins specifically associated with *APOE* or AD, or jointly associated with both. Proteins associated with *APOE* were further tested and categorized based on its upstreaming or downstreaming role in AD using 2 mediation models: *APOE* => protein => AD diagnosis or Aβ status (Upstream mediation model) and *APOE* => AD diagnosis or Aβ status => protein (Downstream mediation model). Diagnosis or Aβ status stratification analysis were conducted to investigate in-depth the changes of *APOE*-protein associations. An age stratification analysis only in CU or Aβ- to investigate how early *APOE*-protein association changes with age. *APOE4* and *APOE2* analysis were conducted separately and the results were compared together with the inclusion of ε2ε4 carriers to investigate protein level change in detailed *APOE* genotypes (6 genotypes: ε2ε2, ε2ε3, ε2ε4, ε3ε3, ε3ε4 and ε4ε4). *APOE* associated proteins were annotated using cell type enrichment analysis, Gene Ontology (GO) term enrichment analysis or biologically informed neural networks (BINNs)-enriched Reactome pathway analysis, protein-protein interaction (PPI) analysis and spatial co-expression analysis (*APOE* and associated genes). Those are done based on the GNPC cohort as it has a larger sample size than other cohorts. For the BioFINDER-2 and ADNI cohort, BINN-enriched reactome analysis was used to globally validate the pathways or processes found in the GNPC cohort. In the BioFINDER-2 cohort, we evaluated the association between *APOE*-associated proteins and available 5 AD biomarkers or cognitive function: tau-PET, Aβ-PET, atrophy: cortical thickness in the temporal lobe, cognitive performance measurements Mini-Mental State Examination (MMSE) and modified Preclinical Alzheimer Cognitive Composite (mPACC) scores. A separate cohort (PPMI) was used to evaluate longitudinal protein-*APOE* associations. Figure created with <u>BioRender.com</u>.

**Fig. 2:**
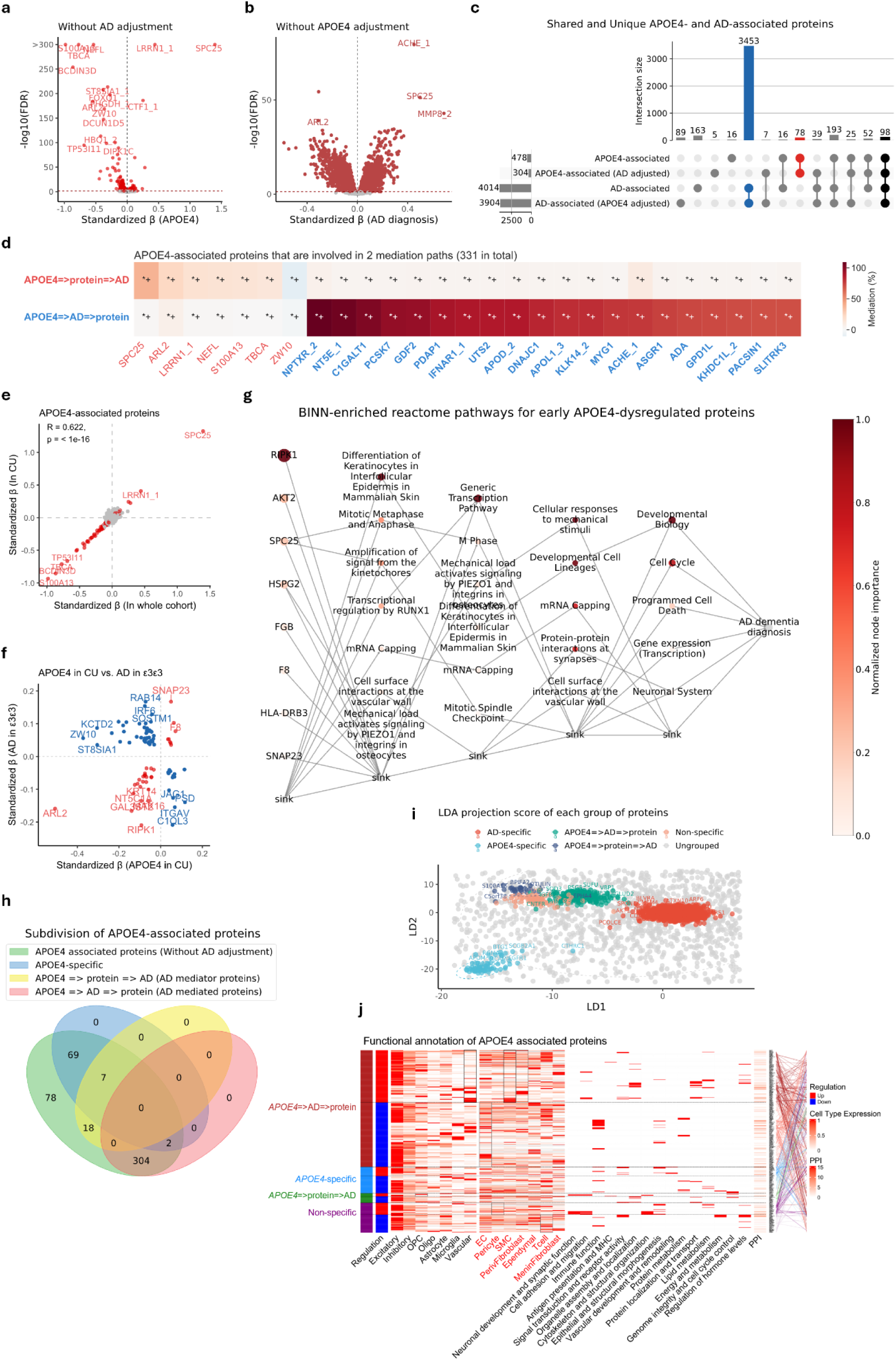
APOE4-associated proteins in GNPC (plasma Somalogic) **a**. Volcano plot for proteins associated with *APOE4* without adjusting for AD diagnosis, with red representing significant association after FDR correction. At y-axis, -log10(FDR) above 300 was set to 300 for a better visualization. **b**. Volcano plot shows proteins associated with AD diagnosis without adjusting for *APOE4*, with red representing significant association after FDR correction. **c**. UpSet plot shows the number of proteins associated with *APOE4* or AD with or without adjusting for each other, blue indicating AD-specific associated proteins, red indicates *APOE4-*specific associated proteins, black indicates the number of proteins independently associated with both. **d**. Heatmap shows the mediation effects and significance of *APOE4-*associated proteins that are involved in the 2 mediation paths. Color indicates the strength of the mediation effect, the darker the color, the bigger the mediation proportion at each path. Proteins at x-axis are categorized and labeled according to their significance in 2 paths (protein-mediated pathway: *APOE4* => protein => AD diagnosis, AD-mediated pathway: *APOE4* => AD diagnosis => protein), with those exhibiting significance in both paths further classified by the strength of the mediation effect. Proteins were marked in red if their protein-mediated pathway was significant alone (after FDR correction) or if the protein-mediated pathway exhibited a stronger mediating effect (when indirect effects in both mediation paths are significant after FDR correction). Proteins were marked in blue if their AD-mediated pathway was significant alone (after FDR correction) or if the AD-mediated pathway exhibited a stronger mediating effect (when indirect effects in both mediation paths are significant after FDR correction). Additionally, bold font indicates a total mediation effect and non-bold font indicates a partial mediation effect in the preferred (same color) path. For heatmap annotations, *represents nominal significant, *+ represents significant after FDR correction. **e**. Scatter plot shows *APOE4*’s effect size on protein without adjusting AD diagnosis in whole cohort (x-axis) vs. in CU individuals (y-axis) for each protein. The Spearman correlation coefficient (R = 0.622, *p* < 1e-16) indicates a moderate positive correlation between the effect sizes. Red represents proteins associated with *APOE4* in both the whole cohort and in the CU group. **f**. The scatter plot shows the effect of *APOE4* on proteins in CU (x-axis) vs. the effect of AD on proteins in ε3ε3 carriers (y-axis); Only proteins associated with *APOE4* in CU individuals and with AD diagnosis in ε3ε3 carriers were visualized. Red indicates the same effect direction while blue indicates an opposite effect direction. **g**. BINN-enriched reactome pathway analysis for proteins associated with *APOE4* in both the whole cohort and in CU. The darker the dot, the more important the protein and the pathway in the deep learning model predicting AD dementia diagnosis. More features are hidden in the sink for a better visualization. **h**. Venn plot shows the number of proteins in each category. Note that 78 proteins were specifically associated proteins in panel **c**, 7 proteins partially mediated the effect of *APOE4* on AD diagnosis, and were therefore subdivided into mediator-proteins group; 2 proteins were partially mediated by AD pathology, and were therefore reassigned into AD-mediated proteins. **i**. The linear discriminant score plot shows the projection score of all tested proteins in the discriminant direction. Proteins were colored by assigned groups. **j**. The integrative matrix summarizes differential regulation (red for upregulated and blue for downregulated proteins in *APOE4* carriers), cell-type enrichment based on scaled RNA expression, and functional characterization of each protein. Cell types from the ROSMAP atlas are labeled in black on the x-axis, while those from the Human Brain Vascular atlas are labeled in red. Gray boxes indicate nominal significance (*p*<0.05), and black boxes indicate FDR-corrected significance (*pFDR*<0.05) in cell-type enrichment analysis. Gene Ontology (GO) biological process terms associated with each protein were grouped into broader representative categories; small red boxes indicate the involvement of a given protein in the corresponding process. Protein–protein interactions (PPIs) are annotated using STRING database interactions with a confidence score≥0.7. The number of interactions per protein is shown as a heatmap, and direct interactions between proteins are represented by lines, color-coded according to their assigned cluster.

#### Mediation reveals bidirectional links between *APOE4*, proteins, and AD diagnosis

To disentangle directionality between *APOE4-*associated proteins and AD, we conducted mediation analyses on the 478 *APOE4*-associated proteins identified in the previous analysis. 25 proteins, e.g., SPC25, ARL2, LRRN1, NEFL, S100A13, TBCA, and ZW10 showed stronger mediation in the *APOE4* => protein => AD pathway than the reverse (Fig. 2d), with partial mediation effects accounting for up to ∼38% of *APOE4*’s total effect on AD risk, consistent with effects upstream of clinical onset (potentially reflecting early pathophysiological changes). In contrast, 306 proteins showed stronger mediation in the *APOE4* => AD => protein pathway, suggesting downstream consequences of disease. 111 proteins, e.g. NPTXR, were affected exclusively through AD (Fig. 2d). For 195 proteins, e.g., RAB14, both direct and AD-mediated effects were observed, with partial AD-mediated (absolute) proportions reaching up to 67% (Supplement Table 2).

#### Effects of *APOE4* in cognitively unimpaired individuals

To explore early, potentially pre-morbid effects of *APOE4*, we performed analysis within CU individuals. *APOE4* was associated with 224 proteins in CU, including 22 of 25 AD-mediators (e.g., SPC25, LRRN1, S100A13, TBCA, Supplement Fig. 1C) and 90 of 195 AD partly-mediated proteins (e.g. RAB14). Most (199, 89%) showed similar significant effect sizes as in the full cohort, supporting *APOE4*’s pre-symptomatic impact (Fig. 2e). Age stratification (median age = 73) revealed that 35% (78) of *APOE4*-associated proteins in CU remained significant in both younger CU and in older CU, including SPC25, TBCA, and S100A13 (Supplement Fig. 1D, E, F). Models including an *APOE4*\*age interaction confirmed largely age-independent effects of *APOE4* on these proteins in CU, except for PPP1R2, WNT10B and DSG2 (Supplement Fig. 1G, H). Among 93 overlapping proteins between *APOE4*-associated alteration in CU and AD-associated alteration in ε3ε3 carriers, *APOE4* had a greater effect on some mediators e.g. ARL2 and ZW10 (Fig. 2f). 31 (e.g., ARL2, SNAP23) showed concordant effects, while 62 showed opposite directions, supporting that *APOE4* may induce early, possibly compensatory changes that differ from those driven by downstream AD pathology.

#### Consistent regulation pattern of *APOE4* across diagnoses

To assess how disease status modifies *APOE4* effects, we also identified *APOE4*-associated proteins within AD cases. Of 123 *APOE4*-associated proteins in AD, 103 overlapped with those in CU and the full cohort, indicating largely diagnosis-independent effects (Supplement Fig. 1I, J). However, two proteins—SPC25 and TBCA—showed significant *APOE4*\*AD interactions: SPC25 was more strongly upregulated by *APOE4* in AD patients, while TBCA exhibited enhanced downregulation (Supplement Fig. 1K, L), potentially reflecting secondary responses to disease progression.

#### Early dysregulated proteins highlight *APOE4*-linked pathways

To elucidate functional consequences of pre-clinical *APOE4*-related proteomic changes, 199 proteins consistently associated with *APOE4* across the full and CU cohorts were mapped to Reactome networks and used to train a deep learning classifier (accuracy: 0.68 training, 0.71 testing) in predicting AD using biologically informed neural networks (BINNs)-based pathway analysis. Key predictors such as RIPK1, AKT2, and SPC25 related cellular responses to mechanical stimuli, developmental cell lineages, mRNA capping, protein-protein interactions at synapses, cell cycle control etc. (Fig. 2g).

#### Stratifying *APOE4*-associated proteins reveals distinct mechanistic routes to AD dementia diagnosis

To further dissect the heterogeneity of *APOE4*-associated proteomic functionality, we subdivided all 478 *APOE4*-associated proteins into four mechanistic categories based on mediation and diagnostic associations (Fig. 2h). Clustering with linear discriminant analysis (LDA) supported biological distinctiveness (Fig. 2i). Cell-type enrichment, pathway, and interaction analyses further supported differences between these four groups of proteins (Fig. 2j):

1. AD mediator proteins (N=25, *APOE4* => protein => AD): Upregulated proteins (N=8, e.g., SPC25 and LRRN1) were enriched in OPCs (nominally) and linked to neuronal development and synaptic function, cell cycle and MHC assembly. Downregulated proteins (N=17, e.g., NEFL, ZW10, TBCA, S100A13 and ARL2) were involved mostly in cytoskeletal organization and organelle localization.
2. AD-mediated proteins (N=306, *APOE4* => AD => protein): Upregulated proteins (N=137) were enriched in vascular cells (e.g., smooth muscle cells and perivascular fibroblasts, the latter with nominal significance) and involved in organelle assembly and localization, protein and hormone regulation. Downregulated proteins (N=169) were mainly linked to immune function.
3. *APOE4*-specific proteins (N=69, associated with *APOE4* but not with AD): Upregulated proteins (N=24) were related to vascular function, tissue remodeling and immune response, while downregulated ones (N=45) involved ubiquitin-dependent catabolic processes and metabolic pathways. 9 proteins were reclassified due to partial mediation effect (Fig. 2h).
4. Non-specific proteins (N=78, associated with both *APOE4* and AD but not involved in either mediation pathway): Upregulated proteins (N=31) were enriched (nominally) in pericytes; downregulated (N=47) in T cells. These proteins are largely clustered together with proteins involved in meditation paths in LDA (Fig. 2i), suggesting overlapping biological patterns.

#### The APOE2-plasma protein signature

Of 211 proteins associated with *APOE2*, 201 (95%) remained significant after adjusting for AD, with 99 specific to *APOE2* without AD association (Supplement Fig. 2A, C). Among 3,055 AD-associated proteins, 3,016 (99%) remained after adjusting for *APOE2*, with 2,911 (97%) specific to AD (Supplement Fig. 2B, C). 88 proteins consistently associated with both *APOE2* and AD. UNG, VPS29 and BIRC2 were most upregulated in *APOE2* carriers and were also upregulated to a similar extent in CU *APOE2* carriers, this including a total of 157 proteins (74% of all 211 *APOE2*-associated proteins) (Fig. 3a, b). Some (N=13), like VPS29 and AKT2, showed an age-dependent pattern with stronger upregulation in younger CU carriers (Fig. 3c, d). Mediation analyses identified 71 proteins (e.g., UNG, ARL2, PCLAF, and SNAP23) for which the *APOE2* => protein => AD diagnosis pathway contributed more than the *APOE2* => AD diagnosis => protein pathway. These proteins partially mediate 5–92% of *APOE2*’s effect on AD. Conversely, 28 proteins (e.g. RAB14) showed partial mediation by AD, with up to 16% effect explained (Supplement Table 2).

**Fig. 3:**
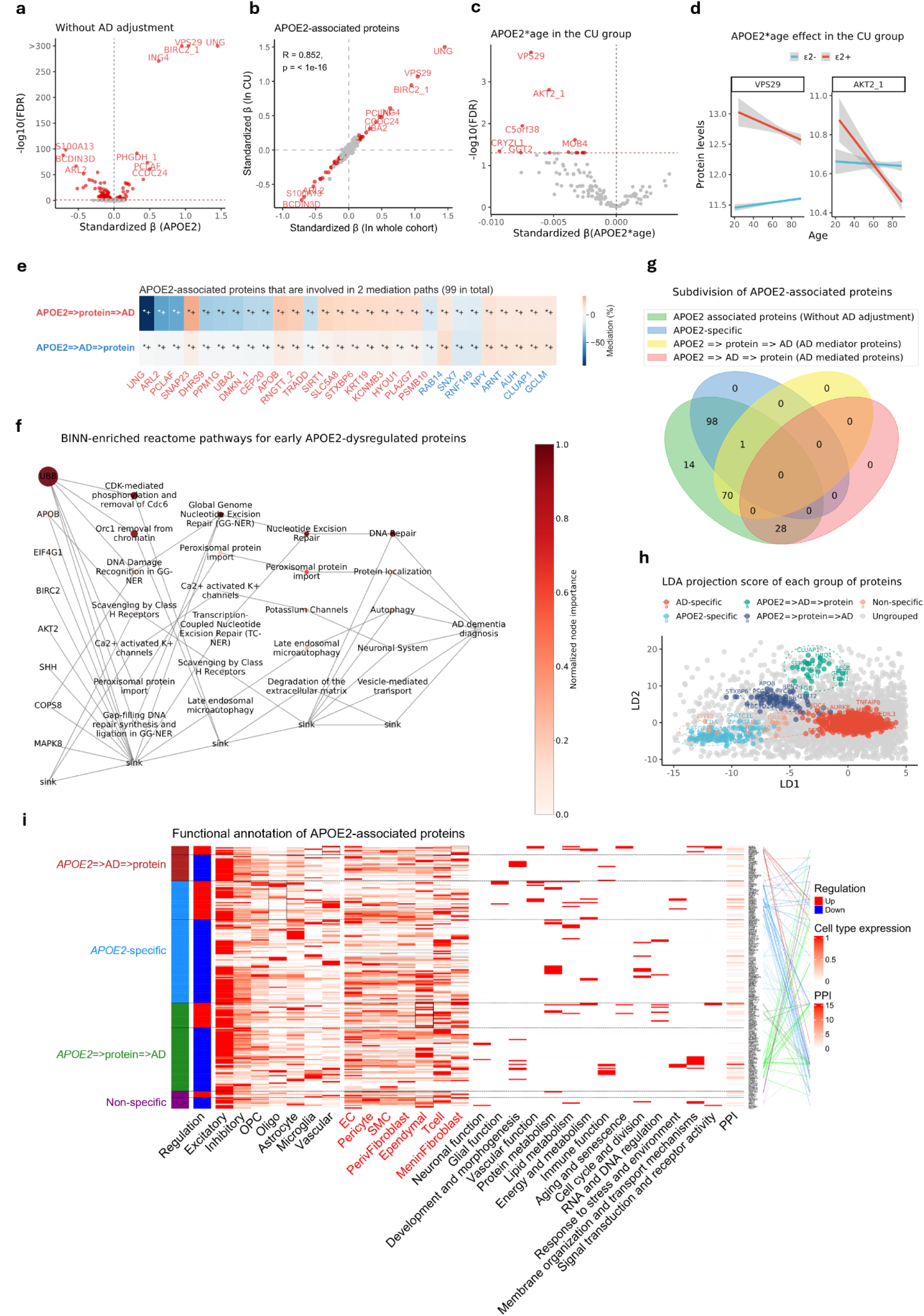
***APOE2* plasma protein signature in GNPC a**. Volcano plot for proteins associated with *APOE2* without adjusting for AD diagnosis, with red representing significant association after FDR correction. At y-axis, -log10(FDR) above 300 was set to 300 for a better visualization. **b**. Scatter plot shows *APOE2*’s effect size on protein without adjusting AD diagnosis in whole cohort (x-axis) vs. in CU individuals (y-axis) for each protein. The Spearman correlation coefficient (R = 0.852, *p* < 1e-16) indicates a strong positive correlation between the effect sizes. Proteins highlighted in red are significantly associated with *APOE2* in both the whole cohort and the CU subgroup. **c**. Volcano plot for the effect of *APOE2*\*age on proteins that are associated with *APOE2* in the CU group with red indicates a significant *APOE2*\*age effect on proteins. **d**. Interaction effect of *APOE2* status and age on protein levels in the CU group. Protein levels of VPS29 and AKT2_1 (“_1” represent one aptamers of AKT2) are plotted against age, with separate regression lines for individuals carrying the ε2 allele (ε2+, red) and those without it (ε2-, blue). Shaded areas represent 95% confidence intervals. **e**. Heatmap shows the mediation effects and significance of *APOE2-*associated proteins that are involved in the 2 mediation paths. Color indicates the strength of the mediation effect, the darker the color, the bigger the mediation proportion at each path. Proteins at x-axis are categorized and colored according to their significance in 2 paths (protein-mediated pathway: *APOE2* => protein => AD diagnosis, AD-mediated pathway: *APOE2* => AD diagnosis => protein), with those exhibiting significance in both paths further classified by the strength of the mediation effect. Proteins were marked in red if their protein-mediated pathway was significant alone (after FDR correction) or if the protein-mediated pathway exhibited a stronger mediating effect (when indirect effects of both mediation paths are significant after FDR correction). Proteins were marked in blue if their AD-mediated pathway was significant alone (after FDR correction) or if the AD-mediated pathway exhibited a stronger mediating effect (when indirect effects of both mediation paths are significant after FDR correction). Additionally, bold font indicates a total mediation effect and non-bold font indicates a partial mediation effect in the preferred (same color) path. For heatmap annotations, *represents nominal significant, *+ represents significant after FDR correction. **f**. BINN-enriched pathway analysis for early dysregulated proteins (associated with *APOE2* in both whole cohort and in CU) in *APOE2* carriers, the darker the color, the more important the protein or pathway in predicting AD dementia diagnosis. More features are hidden in the sink for a better visualization. **g**. Subdivision of *APOE2* associated proteins based on the association between proteins, *APOE2* and AD. Noting that one protein (EMC2) overlaps between *APOE2*-specific and mediator-proteins and was only categorized into mediator proteins for grouped GO biological process enrichment analysis. **h**. The linear discriminant score plot shows the projection score of each group of subdivided proteins in the discriminant direction. Proteins were colored by assigned groups. **i**. The integrative matrix summarizes differential regulation (red for upregulated and blue for downregulated proteins in *APOE2* carriers), cell-type enrichment based on scaled RNA expression, and functional characterization of each protein. Cell types from the ROSMAP atlas are labeled in black on the x-axis, while those from the Human Brain Vascular atlas are labeled in red. Gray boxes indicate nominal significance (*p*<0.05), and black boxes indicate FDR-corrected significance (*pFDR*<0.05) in cell-type enrichment analysis. Gene Ontology (GO) biological process terms associated with each protein were grouped into broader representative categories; small red boxes indicate the involvement of a given protein in the corresponding process. Protein–protein interactions (PPIs) are annotated using STRING database interactions with a confidence score≥0.7. The number of interactions per protein is shown as a heatmap, and direct interactions between proteins are represented by lines, color-coded according to their assigned cluster.

#### Cell-type and functional landscape of *APOE2*-associated proteins

157 consistently identified proteins (full and CU cohorts) were investigated using BINN. The resulting deep learning model (accuracy: 0.74 training, 0.75 testing) revealed top contributors including UBB, APOB, EIF4G1, BIRC2 and AKT2 (Fig. 3f), enriched in DNA repair, peroxisomal protein import and autophagy etc.

We further classified all 211 *APOE2*-associated proteins into four mechanistic categories (Fig. 3g), protein clustering showed a scattered distribution in LDA space (Fig. 3h), indicating distinct functional and spatial features supported by cell type enrichment and subgrouped pathway enrichment results (Fig. 3i, Supplement Table 3):

1. AD mediator proteins (N=71, *APOE2* => protein => AD): Upregulated proteins (N=20, e.g., SIRT1, KMT2C) were enriched in ependymal cells (nominally) and linked to metabolism, chromatin remodeling, and DNA repair; downregulated proteins were tied to membrane organization (SNAP23, APOB etc.), immune and NF-κB pathways (e.g., MAPK8, TRADD), suggesting suppressed inflammation.
2. AD-mediated proteins (N=28, *APOE2* => AD => protein): 7 upregulated proteins, enriched in vascular cells (notably meningeal fibroblasts), supported angiogenesis (e.g., MEOX2).
3. *APOE2*-specific proteins (N=98): Upregulated proteins (N=31, e.g., PHGDH, DLL1) were enriched in oligodendrocytes (nominally) and supported myelination and vascular homeostasis; downregulated ones involved in ubiquitin-mediated proteolysis.
4. Non-specific proteins (N=14): Close to the *APOE2*-specific cluster in LDA space (Fig. 3h), indicating *APOE2*-related functionality. Most (N=9) were downregulated and enriched in ependymal cells (nominally), linked to endomembrane system organization (e.g., ZW10, AKT2).

#### *APOE4* and *APOE2*: Specificity, Offset, and Synergy

*APOE4* and *APOE2* associated proteins were largely non-overlapping (Fig. 4a). Specifically, 8 proteins —including SPC25 and LRRN1—mediated *APOE4*’s effect on AD without being associated with *APOE2*. These proteins showed *APOE4* gene dose–dependent patterns in CU individuals (Fig. 4b), with ε4ε4 showing extreme alteration, and ε2ε4 carriers resembling ε3ε4 carriers (consistent with ε4 dominance). Conversely, 25 proteins (e.g. UNG, PCLAF) mediated *APOE2*’s effect without being associated with *APOE4*, and followed an ε2-dominant pattern (Fig. 4b). 3 proteins (ARL2, OTULIN and TMCC3) mediated both *APOE4* and *APOE2*’s effect on AD diagnosis.

**Fig. 4:**
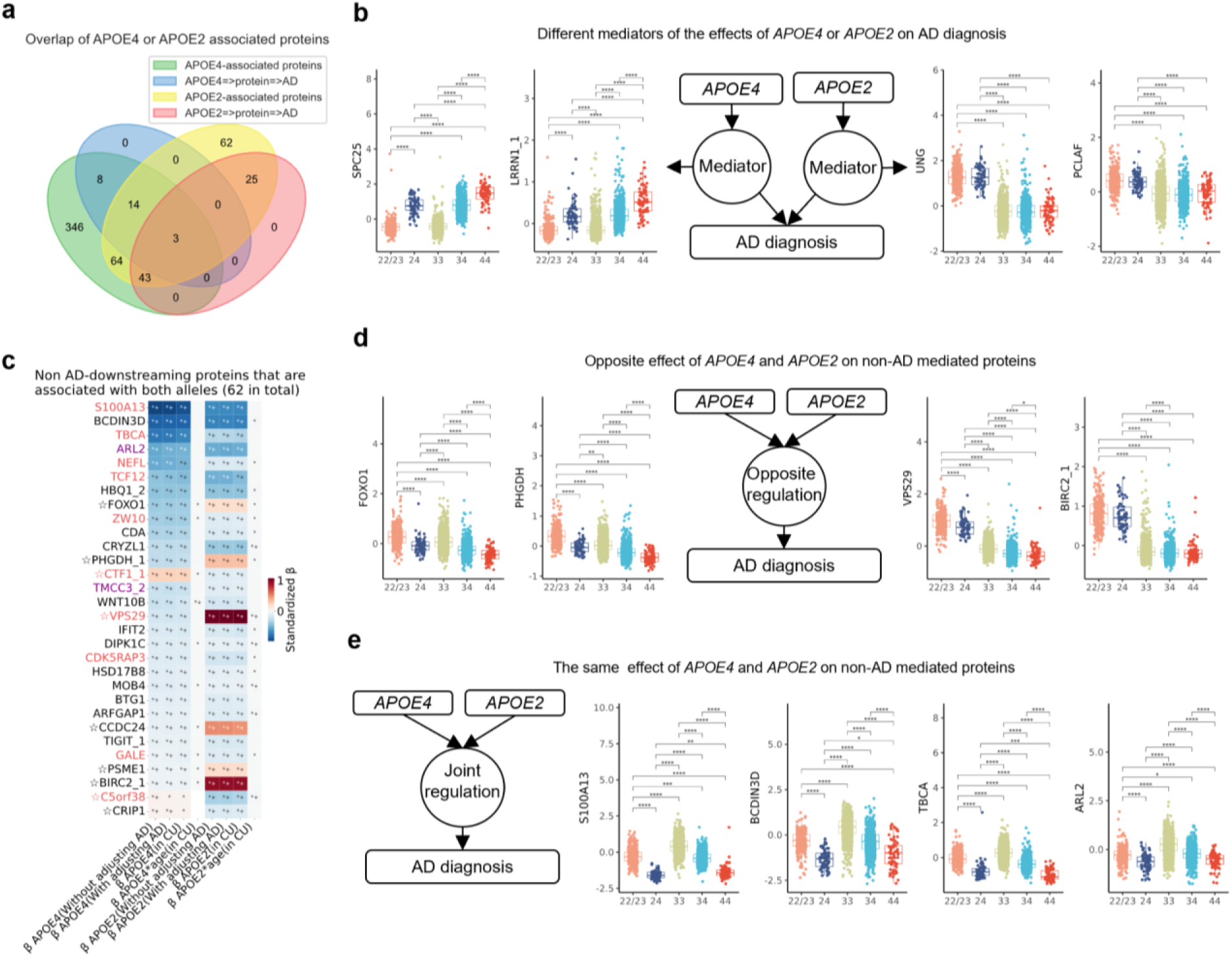
Different pattens of *APOE4*- or *APOE2-*associated proteins. **a**. Venn plot shows proteins mediating *APOE4*’s effect of AD diagnosis and their association with *APOE2*, proteins mediating *APOE2*’s effect on AD diagnosis and their association with *APOE4.* **b**. Boxplots show protein level comparison between different *APOE* genotype groups, for ε4 dominated mediators (SPC25 and LRRN1) and ε2 dominated mediators (UNG and PCLAF). **b. c. d.** On the box plots, the y-axis represents residual protein levels after adjusting for age, sex and mean protein level. The center line of each box indicates the median (50th percentile); the lower and upper edges of the box represent the 25th and 75th percentiles, respectively. Whiskers extend to the most extreme values within 1.5 times the interquartile range (IQR); data points beyond this range are considered outliers and have been excluded from the plot display. The x-axis represents *APOE* genotype, ε2ε3 carriers were merged into the “22/23” group due to a small sample size of ε2ε2 carriers. Welch’s t-test was used to compare residual protein levels between groups. Two-sided p-values are tested and p-values were adjusted for multiple comparisons using Holm-Bonferroni method. Each pair of groups was compared, only pairs with significant differences after FDR correction were shown. Asterisks indicate significance levels: **** for pFDR < 0.0001, *** for pFDR < 0.001, ** for pFDR < 0.01, * for pFDR < 0.05. **c**. Heatmap shows non-AD mediated (non-downstreaming) proteins that are associated with both *APOE4* and *APOE2*, color indicate the effect size of *APOE4* or *APOE2* in each model on those proteins, column 1 indicates *APOE4*’s effect on those proteins without AD diagnosis adjustment, column 2 indicates *APOE4*’s effect on those proteins with AD diagnosis adjustment, column 3 indicates *APOE4*’s effect on those proteins in CU individuals, column 4 indicates the effect of *APOE4*\*age on those proteins in CU individuals.The remaining 4 columns correspond to *APOE2*’s effect: column 5 indicates *APOE2*’s effect on those proteins without AD diagnosis adjustment, column 6 indicates *APOE2*’s effect on those proteins with AD diagnosis adjustment, column 7 indicates *APOE2*’s effect on those proteins in CU individuals, column 8 indicates the effect of *APOE2*\*age on those proteins in CU individuals. “⋆” indicate the opposite effect direction of *APOE4* and *APOE2,* red marked proteins are AD mediator proteins in *APOE4* => protein => AD diagnosis, blue marked proteins are AD mediator proteins in *APOE2* => protein => AD diagnosis, purple marked proteins are AD mediator proteins in both. Only proteins for which the average of the absolute value of the effect of *APOE4* and *APOE2* were greater than 0.1 are shown. For heatmap annotations, *represents nominal significant, *+ represents significant after FDR correction. **d**. Boxplots show protein level comparison between different *APOE* genotype groups for non-downstreaming proteins that are opposingly regulated by both alleles. **e**. Boxplots show protein level comparison between different *APOE* genotype groups for non-downstreaming proteins that are similarly regulated by both alleles.

SNRPF mediated *APOE4*’s effect on AD but appeared downstream of pathology in *APOE2* models, while 41 proteins—including SNAP23 and APOB—showed the opposite pattern (Supplement Table 2). These shifts underscore divergent molecular pathways for *APOE4* and *APOE2* in AD. 62 proteins were not mediated by AD in either *APOE4* or *APOE2* models— suggesting upstream regulation—yet were associated with both alleles. 13 showed opposing allele effects in CU individuals (Fig. 4c), following either ε4-dominant (e.g., FOXO1, PHGDH) or ε2-dominant (e.g., VPS29, BIRC2) patterns (Fig. 4d). The remaining 49 proteins were similarly regulated by both alleles; key mediators such as S100A13, BCDIN3D, TBCA, and ARL2 were downregulated in both ε2 and ε4 carriers, with ε2ε4 individuals resembling ε4ε4 carriers (Fig. 4e).

#### *APOE4*, plasma Proteins, and Aβ pathology: BioFINDER-2 Cohort The BioFINDER-2 plasma SomaLogic Cohort

BioFINDER-2 cohort is a subset of the GNPC with deeper phenotyping, where whether plasma proteins mediated the effects of *APOE* on Aβ pathology was tested. The same proteins as in GNPC were available for testing (Supplement Fig. 3A). Focusing on Aβ pathology, 259 AD dementia, 316 Mild Cognitive Impairment (MCI) and 846 CU participants were included, resulting in 1,421 participants in total (aged 20–93 years; median age: 72). Based on CSF Aβ42/40 ratios, 715 individuals (50.3%) were Aβ- and 706 (49.7%) were Aβ+ (Supplement Table 1).

#### Mediators of clinical AD vs. Aβ pathology

Consistent with GNPC findings, most plasma proteins altered in *APOE4* carriers were independent of Aβ status but showed Aβ-mediated changes. 65 proteins (39% of *APOE4*-associated proteins identified in this cohort) were altered in both the full cohort and Aβ-individuals, supporting early, preclinical effects. Among them, AKT2, HDAC8, APOB, CYP3A4, SPC25, and BCDIN3D—linked to chromatin organization, gene expression, drug ADME and cell cycle—were top features in the BINN model predicting Aβ status in CU and MCI individuals (Supplement Fig. 4A).

Similarly, 43 (59% of *APOE2*-associated proteins) proteins were already altered in Aβ-*APOE2* carriers. Among these, key Aβ predictors like APOB, SHH, and BIRC2 were enriched in pathways like mitochondrial import, SLC transport, lipid metabolism, and adaptive immunity (Supplement Fig. 4B).

Mediation analysis identified 8 proteins (e.g., SPC25, TBCA, S100A13, CTF1, BCDIN3D) partially mediated *APOE4*’s effect on Aβ pathology (4–35%; Fig. 5a); 7 remained significant after adjusting for population stratification (except UBL3; Supplement Fig. 5A). Ten proteins mediated *APOE4*’s effect on AD diagnosis (5–83%) (Fig. 5b), among these, SPC25, TBCA, and S100A13 replicated from GNPC (Fig. 2d) and showed full mediation, 5 proteins, including S100A13, TBCA, and PHGDH, remained robust after adjusting for population stratification (Supplement Fig. 5B).

**Fig. 5:**
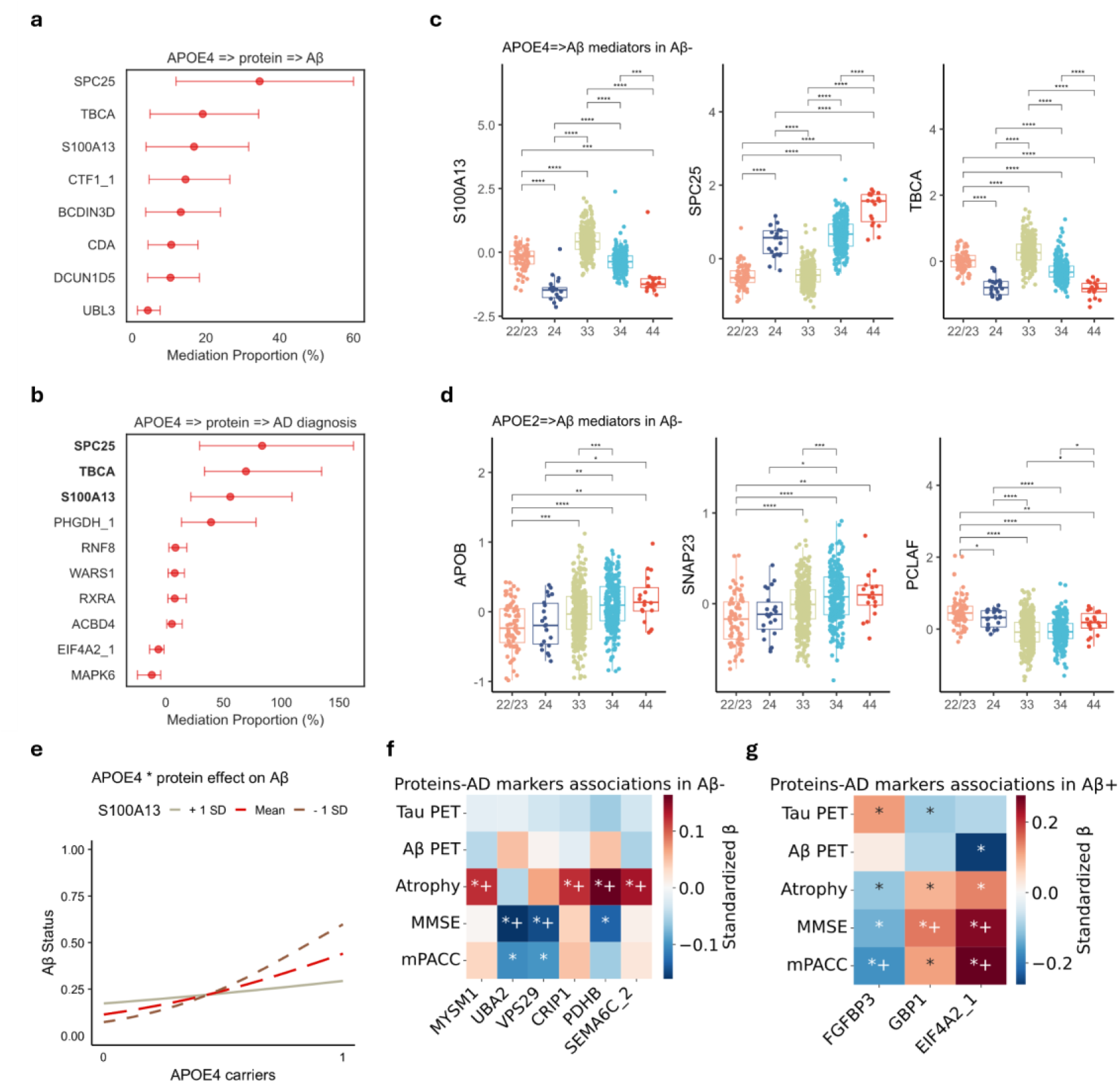
Mediators of clinical AD vs. AB pathology (BioFINDER-2 plasma Somalogic) **a**. The dot plot with error bars shows the mediation proportions and confidence intervals for proteins mediating the effects of *APOE4* on Aβ. **b**. The same but for proteins mediating the effects of *APOE4* on AD diagnosis. For similar plots **a**, **b**, proteins whose indirect effects in protein mediation path are significant after FDR correction alone or mediation proportions of protein-mediation pathways are greater than those of Aβ (or AD) -mediated pathways (when indirect effects of both mediation paths are significant after FDR correction) are defined as mediators and are shown. The x-axis represents the percentage of mediation proportions. The dots represent the estimated mediation proportions of each protein while red indicates a significant estimation. The horizontal lines represent the 95% confidence intervals of these estimates. Bold indicates that the direct effect of *APOE4* is not significant, resulting in a total mediation effect of this protein. **c**. Boxplots show protein level change groups by *APOE* genotypes for key *APOE4* => Aβ mediators in Aβ-individuals. **d**. The same but for key *APOE2* => Aβ mediators in Aβ-individuals. On the box plots **c** and **d**, the y-axis represents residual protein levels after adjusting for age, sex and mean protein level. The center line of each box indicates the median (50th percentile); the lower and upper edges of the box represent the 25th and 75th percentiles, respectively. Whiskers extend to the most extreme values within 1.5 times the interquartile range (IQR); data points beyond this range are considered outliers and have been excluded from the plot display. The x-axis represents *APOE* genotype, ε2ε2 and ε2ε3 carriers were merged into the “22/23” group due to a small sample size of ε2ε2 carriers. Welch’s t-test was used to compare residual protein levels between groups. Two-sided p-values are tested, and p-values were adjusted for multiple comparisons using Holm-Bonferroni method. Each pair of groups was compared, only pairs with significant differences after FDR correction were shown. Asterisks indicate significance levels: **** for pFDR < 0.0001, *** for pFDR < 0.001, ** for pFDR < 0.01, * for pFDR < 0.05. **e**. The interaction plot shows the interaction between *APOE4* carrier status (0 = non-carriers [ε3ε3], 1 = carriers) and S100A13 protein expression in predicting Aβ status in CU and MCI groups. Lines represent estimated Aβ status across *APOE4* groups at three levels of S100A13 expression: one standard deviation below the mean (−1 SD, dashed gray), the mean (solid red), and one standard deviation above the mean (+1 SD, dashed brown). Among *APOE4* carriers, higher S100A13 levels are associated with a greater likelihood of elevated Aβ status. **f**. The heatmap displays standardized regression coefficients (β) from models examining the associations between *APOE*-associated proteins (x-axis) and AD-related markers (tau-PET: tau-PET uptake in the temporal meta-ROI, Aβ-PET, Atrophy: cortical thickness in the temporal lobe, MMSE and mPACC: the modified preclinical Alzheimer’s cognitive composite) (y-axis) in Aβ-individuals. Full statistical details are available in Supplement Table 7. **g**. The same but in Aβ+ individuals. On heatmaps, red shades indicate positive associations; blue shades indicate negative associations. The darker the color, the stronger the associations between *APOE*-associated proteins and 5 AD biomarkers or cognitive performance. For heatmap annotation, * indicates a nominally significant association, *+ indicates a significant association after FDR correction.

Most *APOE2*-associated proteins were independent of Aβ and showed no mediation effect by Aβ. However, APOB, SNAP23, WARS2, and PCLAF—replicated from GNPC—fully mediated *APOE2*’s effect on Aβ pathology (Supplement Table 2). No mediators were identified for *APOE2*’s effect on clinical AD diagnosis.

Alterations of these mediators were detectable in Aβ-individuals, largely age-independent (except SPC25), and showed allele-specific patterns: *APOE4* mediators displayed ε4 dose-dependence and ε4-dominance in ε2ε4 carriers, while *APOE2* mediators showed ε2-dominant effects (Fig. 5c, d).

#### *APOE4*\*protein interaction effects on Aβ status

Beyond mediation, we assessed whether *APOE4*-associated proteins modulate genetic risk. An *APOE4*\*protein interaction analysis revealed that S100A13 significantly influenced the relationship between *APOE4* and Aβ status: *APOE4* carriers with lower S100A13 levels showed higher Aβ positivity in CU and MCI individuals, an effect diminished at higher protein levels (Fig. 5e).

#### Associations between *APOE*-related proteins and AD features in BioFINDER-2

To evaluate functional relevance, we assessed associations between *APOE*-related proteins and AD features (tau-PET, cortical thickness, cognition) in Aβ- (N=715) and Aβ+ (N=706) individuals. In the Aβ-group, higher VPS29 and UBA2 levels were associated with worse cognition, lower MYSM1, CRIP1, PDHB or SEMA6C levels were associated with more severe atrophy (Fig. 5f). In Aβ+, lower FGFBP3, higher EIF4A2 or GBP1 levels were associated with better cognition (Fig. 5g). Those proteins are either *APOE* (*APOE4* or *APOE2*)-specific or associated with both *APOE* and Aβ with no mediation effect. The lack of associations between mediator proteins and AD biomarkers or cognition may reflect their early role in *APOE*-related pathways, prior to the emergence of downstream pathology and clinical symptoms.

#### CSF proteomic signature in SomaLogic proteomics (ADNI cohort)

In ADNI CSF (N=666; SomaLogic 7k; aged 54-91, median age 74; 58% Aβ+; 325 ε4, 46 ε2, 295 ε3ε3 carriers), we validated *APOE*-associated pathways and mediators. *APOE4* was associated with 684 proteins (124 GNPC-consistent), 48% independent of Aβ status; 122 already altered in Aβ-individuals, including BINN top features like AKT2 and ARF1 (accuracy=0.66 for training and testing), enriched in ECM, autophagy, meiosis and RNA pathways (Supplement Fig 6A).

*APOE2* was associated with 93 proteins (all evident in Aβ-, 35 GNPC-consistent), non-overlapping with Aβ-related proteins; BINN predictors (accuracy: 0.70 training, 0.66 testing) such as PTPN1 and MAPK8 were enriched in IGF signaling, autophagy, and DNA repair (Supplement Fig 6B).

Mediation analysis identified 20 proteins linking *APOE4* to Aβ (2–57% mediation; Fig. 6a) while more proteins (251) were downstream of Aβ (Supplement Table 2). Several mediators overlapped with plasma results (Fig. 6b): S100A13 and TBCA were consistent across GNPC and BioFINDER-2; BCDIN3D was consistent when using Aβ as outcome, indicating Aβ-specific mediation; CSF SPC25 was not a mediator despite its plasma effect, suggesting matrix specificity. No *APOE2* mediators were identified (our power was very limited here due to few ε2 carriers in ADNI).

**Fig. 6:**
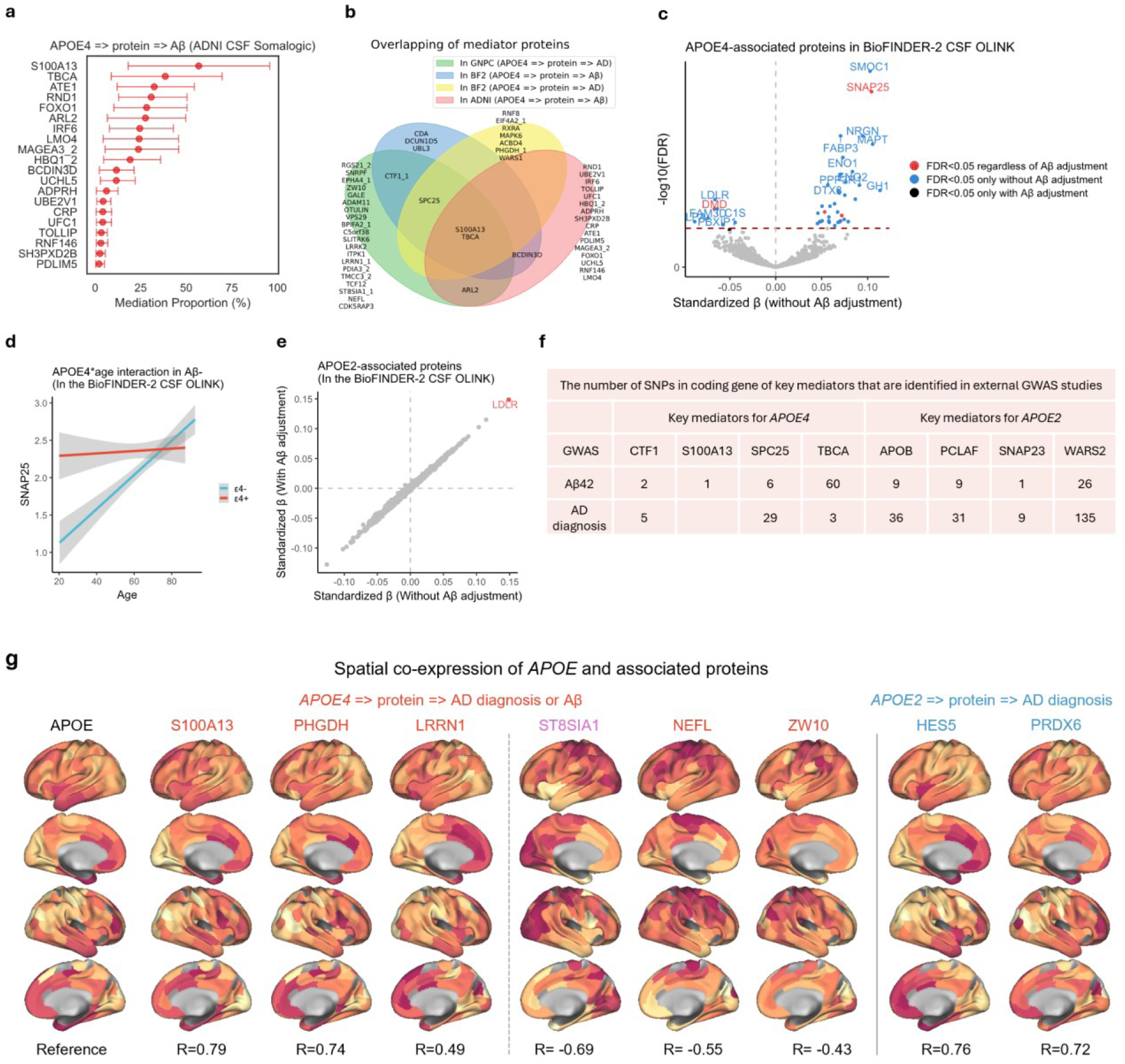
Validation in CSF proteomics, GWAS and transcriptome. **a**. The dot plot with error bars shows the mediation proportions and confidence intervals for proteins mediating the effects of *APOE4* on Aβ in ADNI. Proteins whose indirect effects in protein mediation path are significant after FDR alone or mediation proportions of protein-mediation pathways are greater than those of Aβ (or AD) -mediated pathways (when indirect effects of both mediation paths are significant after FDR) are defined as mediators and are shown. The x-axis represents the percentage of mediation proportions. The dots represent the estimated mediation proportions of each protein while red indicates a significant estimation. The horizontal lines represent the 95% confidence intervals of these estimates. **b**. Venn plot shows mediators identified in 3 SomaLogic proteomics dataset. **c**. Volcano plot shows *APOE4* associated proteins with or without Aβ adjustment in the BioFINDER-2 CSF OLINK cohort, x-axis is the effect size of *APOE4* on proteins without adjusting Aβ, red represent proteins associated with *APOE4* regardless of adjusting Aβ status while blue indicate proteins associated with *APOE4* only without adjusting Aβ. **d**. Stratified linear fit plot shows SNAP25 protein level change with age in *APOE4* carriers and non-carriers in Aβ-individuals in the BioFINDER-2 CSF OLINK cohort. SNAP25 levels are plotted against age, with separate regression lines for individuals carrying the ε4 allele (ε4+, red) and those without it (ε4-, blue). Shaded areas represent 95% confidence intervals. **e**. Scatter plot shows proteins associated with *APOE2* with or without adjusting Aβ status in the BioFINDER-2 CSF OLINK cohort. The red dot indicates a significant association regardless of adjusting Aβ. **f**. The table shows the number of SNPs in the coding gene of key mediators (identified as mediators in at least 2 dataset) that are associated with AD clinical diagnosis or CSF Aβ42 level in external GWAS studies. **g**. The plot shows brain-wide gene expression patterns, with statistics reflecting spatial correlations between *APOE* and *APOE4* or *APOE2* associated proteins. Only key proteins with significant spatial coexpression with *APOE* were shown. Red indicates mediators of the *APOE4* => protein => AD diagnosis or Aβ pathology pathway; blue denotes mediators of the *APOE2* => protein => AD diagnosis pathway. Purple marks proteins implicated in both, we note that ST8SIA1 were identified as mediators in the 2 mediation pathways by two distinct aptamers (seq.21508.7 for *APOE4*, seq.21663.149 for *APOE2*)

#### CSF proteomic signature in OLINK proteomics (BioFINDER-2 cohort)

To validate CSF *APOE* proteomic findings in a different proteomic platform, we utilized OLINK proteomics data in the BioFINDER-2 cohort (N=1,475, 1,391 proteins, several key mediators e.g., SPC25, CTF1, ARL2, ZW10, APOB, PCLAF and WARS2 were unavailable). Available key mediators e.g., S100A13, TBCA and SNAP23 were not significantly associated with *APOE* in CSF OLINK. Of 43 *APOE4*-associated proteins, most lost significance after Aβ adjustment, except for 5 proteins e.g., SNAP25 and DMD (Fig. 6c). Mediation analysis showed weak *APOE4* => Aβ partial mediation via CKAP4 (2%) and partial *APOE4* => AD mediation via SNAP25 (7%) (Supplement Table 2). SNAP25 was more upregulated in young Aβ-*APOE4* carriers, with age-dependent attenuation (Fig. 6d). For *APOE2*, only LDLR showed significant association regardless of Aβ adjustment (Fig. 6e) but did not mediate AD diagnosis or Aβ pathology.

#### Plasma proteomic signature in OLINK proteomics (UKB cohort)

To validate plasma *APOE* proteomic signatures in a different proteomic platform, we utilized the UKB cohort (N= 4,820, 1,319 OLINK proteins, most key mediators from the SomaLogic analyses were unavailable in the OLINK assays). 10 *APOE4* and 38 *APOE2* associated proteins were identified, among these, BRK1 and PLA2G7 were associated with both alleles in opposing directions (Supplementary Fig. 7A, B). Interestingly, both *APOE4* and *APOE2* showed gene-dose effects on PLA2G7 (Supplementary Fig. 7C). LDLR was markedly elevated in ε2ε2 individuals (Supplement Fig. 7C).

#### SNPs associated with Aβ42 measurements and AD in coding genes of mediator-proteins

In external GWAS data from the the GCST002245 study^28^, of the 33 proteins identified as mediators of *APOE4*’s effect on AD in GNPC, BioFINDER-2 or ADNI, 20 had coding genes harboring AD-associated SNPs. Similarly, among 71 *APOE2* => AD mediator proteins, 35 had AD-associated SNPs. Based on the GCST90129599 study^29^, 17 of 26 *APOE4* => Aβ mediators and all 4 *APOE2* => Aβ mediators had SNPs associated with CSF Aβ42 levels. Notably, among the 6 key proteins mediating *APOE4*’s effect on AD diagnosis or Aβ pathology in at least two datasets (Fig. 6f), 4 (S100A13, SPC25, TBCA and CTF1) had coding variants linked to Aβ42 or AD. All 4 key *APOE2* mediators (APOB, PCLAF, SNAP23, WARS2) identified in multiple datasets were also supported by SNP associations with Aβ42 or AD (Fig. 6f).

#### Spatial Overlap Analyses

*APOE*-protein spatial co-expression was validated using Allen Human Brain Atlas transcriptomic data. Among 368 *APOE4*-associated proteins (in GNPC) with available expression data, 170 (46%) showed significant spatial correlation with *APOE*, including 10 AD or Aβ mediators e.g. positive associations for S100A13, PHGDH, and LRRN1, and negative associations for ST8SIA1, NEFL, and ZW10 (Fig. 4f). Of 166 *APOE2*-associated proteins, 70 (42%) showed spatial correlation with *APOE*, including 21 AD mediators such as HES5 (R = 0.76) and PRDX6 (R = 0.72). Several key mediators, e.g., SPC25, APOB, and PCLAF, were unavailable for transcriptomic testing. No spatial correlation was observed between Aβ-PET signal and the expression of *APOE* or any *APOE*-associated genes (Supplement Table 4).

#### Dynamic profiling in CSF supports APOE temporal effects

To investigate longitudinal *APOE*-protein associations, we utilized the PPMI CSF OLINK cohort (N=253), among 72 *APOE4* and 12 *APOE2* associated proteins from GNPC, only LGALS9, PRSS8 and NEFL showed nominally significant *APOE*\*time interactions. Similarly, among 26 testable proteins from BioFINDER-2 CSF OLINK, only SIGLEC1 showed a nominally significant *APOE4*\*time interaction. Suggesting that *APOE*-associated proteomic signatures are largely stable over time, supporting their role as early and persistent molecular features rather than dynamic markers of progression. Full description is in Supplement text.

#### Cross-platform and cross-tissue variability of APOE-associated proteomic signatures

*APOE4*-associated protein profiles showed strong consistency within the same tissue and platform (e.g., r=0.72 between plasma SomaLogic datasets), but substantially lower correlations across tissues and platforms (Supplement Fig. 8A-E). NEFL exhibited notable heterogeneity across platforms (Supplement Fig. 8F, G). No protein was consistently associated with *APOE4* or *APOE2* across all datasets and technologies, indicating high heterozygosity of proteomics findings across tissues or platforms (Supplement Fig. 8H, I). Full description is in Supplement text.

## Discussion

We conducted a large-scale, cross-platform proteomic analysis of plasma and CSF from over 10,000 samples to map how *APOE* isoforms (ε2, ε4) modulate AD risk. *APOE*-associated proteins were classified into risk mediators, protective mediators, downstream reflectors, and neutral markers, revealing largely distinct pathways for *APOE4* and *APOE2*.

Key *APOE4*-associated mediators—such as SPC25 (involved in kinetochore assembly, previously linked to microglial activation in mouse models^30^ and elevated in MCI patients^31^), S100A13 (moderating Aβ positivity in *APOE4* carriers, and implicated in cellular senescence^32^), and ZW10 (a spindle checkpoint protein), some were enriched in OPCs—point to dysregulation of cell cycle control and mitotic processes as potential contributors to AD risk and Aβ accumulation^33–36^, especially in glial cells. TBCA and ARL2 further highlight cytoskeletal disruption as an *APOE4*-related feature contributing to neurodegeneration^37,38^. In contrast, *APOE2*-associated mediators—including SNAP23 (endocytic trafficking), APOB (suggesting altered lipid transport and metabolism), WARS2 (mitochondrial translation), and PCLAF (DNA repair)—suggest a protective effect of *APOE2* via maintenance of cellular homeostasis, mitochondrial function, and genomic stability^39–42^. These mechanistic insights are supported by consistent signals across datasets, reinforced by GWAS and gene-dosage associations.

Together, they nominate distinct, *APOE* genotype-specific biological processes—particularly those involving cell cycle regulation, cytoskeletal integrity, and mitochondrial health—as promising therapeutic targets for AD.

While individual mediators reveal discrete mechanisms, our broader analysis reveals a temporally dynamic *APOE4*-driven program: asymptomatic *APOE4* carriers exhibit early proteomic changes implicating cell cycle, stress responses, and cytoskeletal disruption, preceding amyloid or symptoms. As pathology advances, more *APOE4*-associated proteins become AD-mediated, including vascular proteins involved in organelle organization and adhesion, alongside downregulated immune-related proteins^43–45^, reflecting a staged progression from genetic priming to pathology-driven cascades.

Beyond merely countering *APOE4*^46–48^, *APOE2* confers protection through early molecular programs—such as DNA repair, vesicular trafficking, and mitochondrial function— detectable even in CU or Aβ-individuals and supported by prior studies^49–51^. Most *APOE2* proteins act as upstream of AD with non-Aβ mediated effect and minimal mediation by AD, suggesting early buffering of pathology (may explain better baseline cognitive ability in *APOE2* carriers^52^) that may wane over time. Functional classification delineates four categories: (1) AD mediators promoting metabolism, DNA repair, and reduced inflammation; (2) AD-mediated proteins involved in vascular remodeling and angiogenesis aligning with AD-mediated proteins in *APOE4* carriers; (3) *APOE2*-specific proteins supporting oligodendrocyte function, myelination, and proteostasis; and (4) non-specific proteins linked to endomembrane and ependymal function—together defining a comprehensive, system-level functional classification of *APOE2*-associated pathways, addressing a critical gap in the current understanding of *APOE* isoform biology.

Though largely distinct, some proteins are dysregulated by both alleles, showing allele-specific dominance patterns—e.g., FOXO1 and PHGDH (*APOE4* dominant), VPS29 and BIRC2 (*APOE2* dominant)—indicating genotype-specific molecular switches. Some proteins (ARL2, TBCA, S100A13) are similarly affected by both, with *APOE2* occasionally mimicking ε4-like effects, which may partly explain reports linking *APOE2* to neurodegenerative risk under certain conditions^52^.

Strengths of our study include the integration of multiple platforms and tissues across >10,000 participants, enabling a comprehensive *APOE* molecular map. Mediation modeling separates causal mediators from downstream cascades, providing mechanistic insight beyond correlations. Key results were robustly replicated and supported by genetic and transcriptomic data, although variability across datasets highlights the need for context-aware analyses and validation^53,54^. Limitations include platform-related protein coverage differences, limited ε2 carrier numbers reducing power especially in CSF and longitudinal analyses, and a largely cross-sectional design limiting temporal resolution. Causality inferred by statistical mediation requires functional validation.

In summary, our integrative proteomic analysis reveals that *APOE4* and *APOE2* regulate AD risk through distinct, largely non-overlapping pathways: *APOE4* induces early cell cycle dysfunction, especially in glial cells, while *APOE2* enhances resilience through DNA repair, inflammation reduction and mitochondrial function. These allele-specific changes appear before symptoms or Aβ pathology, highlighting a critical early intervention window. The consistency of these findings across diverse platforms and tissues highlights robust, genotype-specific biomarkers and intervention targets, based on *APOE*-linked proteomic changes.

## Methods

### GNPC SomaLogic 7K participants

The GNPC is a multi-center, international proteomics initiative. The cohort composition, sample processing, and proteomics pipeline have been described in detail previously^55^. The present study represents a focused deep dive into the GNPC dataset to investigate the role of *APOE4* and *APOE2* in AD, using the same inclusion criteria and preprocessing steps as in Imam et al. (2025).

In total, 4,045 participants were included (aged 20–90 years; median age 75), comprising 2,566 CU individuals and 1,479 patients with AD dementia. Plasma proteomics were profiled using the SomaScan® 7K platform (SomaLogic Inc.), which quantifies proteins with DNA-based SOMAmer® reagents^56^. Proteomics data underwent the standard SomaScan® pipeline, including quality control and adaptive normalization by maximum likelihood (ANML).

*APOE*-targeting aptamers were excluded, resulting in 7,285 aptamers corresponding to 6,358 unique proteins. All aptamer intensities were log2-transformed, and values deviating more than 5 standard deviations from the mean were defined as outliers and excluded. This preprocessing was applied to all SomaLogic proteomics datasets used in this study.

For primary analyses, ε2ε4 carriers were excluded to avoid confounding^55^. *APOE4* analyses compared ε3ε4 and ε4ε4 carriers to ε3ε3 (binary variable), and *APOE2* analyses compared ε2ε2 and ε2ε3 carriers to ε3ε3. ε2ε4 carriers were only included in descriptive comparisons of all six genotype groups (ε2ε2, ε2ε3, ε2ε4, ε3ε3, ε3ε4, ε4ε4), consistent with GNPC, BioFINDER-2, and UK Biobank analyses.

### BioFINDER-2 cohort

The Swedish BioFINDER-2 study (NCT03174938; https://biofinder.se) was one of the data contribution sites in GNPC with available detailed AD phenotyping data, e.g., Aβ data, including CSF Aβ42/Aβ40 ratio and Aβ PET. We used BioFINDER-2 plasma SomaLogic data to validate the plasma proteomics of *APOE* (*APOE4* and *APOE2*) and explore the role of these proteins in *APOE*-Aβ associations. Focused on Aβ-related pathological changes across AD progression, we included 846 CU, 316 MCI, and 259 AD individuals with genotyping and SomaLogic proteomics measurements in plasma, resulting in 1,421 individuals (aged 20-93, median age 72) in total. Other dementia types that might introduce confounding neurodegenerative pathways were excluded. The details of these sub-cohorts have been described previously, including the diagnostic criteria^57^ and cognitive staging^58^. Aβ status (positive or negative) were defined using CSF Aβ42/Aβ40 ratio (CSF Aβ42 and Aβ40 were measured using the Roche Diagnostics Elecsys® CSF electrochemiluminescence immunoassay and Roche NeuroToolKit respectively) as described before^59–61^. The initial processing of proteomics data was the same as the GNPC dataset.

We also explored and compared *APOE*-associated CSF proteins and Aβ effects on them using CSF proteomics data from OLINK Explore 3072 proximity extension assay. A total of 1,567 individuals (aged 20-93, median age 72) were included. Like the plasma SomaLogic dataset, we only included CU and subjects diagnosed with MCI and AD, excluding individuals with a suspected neurodegenerative disease other than AD. Protein quality control was conducted as described^22^. Specifically, the normalized protein expression NPX value of each protein was compared to its Limit Of Detection (LOD), and only proteins with levels above the LOD in at least 70% of participants were retained. APOE-proteins were also excluded. Resulting in 1,391 measurements (tested and reported as independent proteins as in all cohorts. 1,382 unique proteins, Supplement Text and Figures) in total. The mean protein level for 1,391 proteins was calculated for each included subject and used as a covariate in all models^62^.

In order to investigate the heterozygosity of protein measurements across multiple platforms, in the BioFINDER-2 cohort, plasma Nfl measured using the Quanterix Simoa® NfL assay, CSF NfL level measured using Roche Elecsys NeuroToolKit (NTK) assay were utilized and were compared to plasma SomaLogic and CSF OLINK measurements.

Ethically, written informed consent was obtained from all participants prior to entering the study. Ethical approval was obtained from the Regional Ethical Committee in Lund, Sweden.

### ADNI

*APOE*-protein profiles in CSF and proteins mediate *APOE*’s effect on Aβ were validated in ADNI. ADNI is a multisite, longitudinal study launched in 2003 as a public-private partnership, with the primary aim of determining whether serial MRI, PET imaging, fluid biomarkers, and clinical/neuropsychological assessments can be integrated to track the progression of MCI and early AD. Further details about the study are available at www.adni-info.org. A total of 666 CU, MCI, AD dementia individuals with available *APOE* genotyping and CSF proteomic profiles measured using the SomaScan 7K platform (v4.1) were included. Aβ status was defined using Aβ-PET, as previously described^63^. The *APOE* protein itself was excluded from downstream analyses, resulting in a final dataset comprising 7,001 quantified proteins. SomaLogic proteomics data were processed similarly as described above.

### UK Biobank

In order to validate and compare *APOE*-associated protein profiles across tissues and proteomics platforms, *APOE*-associated proteins were identified in data from the UK Biobank dataset^64^. This large-scale, multicenter prospective cohort study included approximately 500,000 participants aged 40 to 69 years, who were enrolled between 2006 and 2010. For more information on study methods and data collection, see the UKB online protocol (www.ukbiobank.ac.uk). 4,820 participants were included, all of whom had genotype data and protein levels in plasma measured using OLINK Explore 3072. The same protein retention criteria as for BioFINDER-2 OLINK dataset were applied and 1,319 proteins were ultimately included.

### PPMI

To explore longitudinal *APOE*-protein association, we utilized data from the Parkinson’s Progression Markers Initiative (PPMI), which is an ongoing, international, observational study conducted across the United States, Europe, Israel, and Australia. To date, the study has enrolled approximately 4,000 participants, including healthy controls (HC), de novo Parkinson’s Disease (PD) patients, prodromal individuals (aged 60 or older with a dopamine transporter [DAT] deficit and either REM sleep behavior disorder [RBD] or hyposmia), and non-manifesting carriers of LRRK2 and GBA gene mutations. Participants undergo in-depth clinical assessments, neuroimaging, and molecular phenotyping. The study was approved by the Institutional Review Board at each site, and participants provided written informed consent.

For this study, we utilized the curated dataset from the PPMI sub-study Project 9000. Briefly, Project 9000 includes PD patients, prodromal individuals, and healthy control participants. Longitudinal proteomics profiling was performed on CSF samples using the OLINK Explore platform. A full study protocol is available at: https://ida.loni.usc.edu/download/files.

260 participants with available CSF proteomics data were included, including 74 PD patients, 56 prodromal individuals, and 130 healthy controls. Only proteins for which at least 70% of participants had levels above the LOD were retained as described above, resulting in 826 proteins. Additionally, the mean overall protein levels were also calculated and included as a nuisance covariate in all models.

### Main statistical models

Proteins associated with *APOE4* allele carrier status (binary, carriers [ε3ε4 and ε4ε4] vs. ε3ε3 carriers) were analyzed using linear model 1. Proteins associated with AD diagnosis (binary, CU vs. AD) in GNPC or with Aβ status (binary, Aβ- vs. Aβ+) defined by CSF Aβ42/Aβ40 ratio in the BioFINDER-2 cohort and by Aβ-PET in the ADNI cohort were analyzed using linear model 2. A third model incorporated both *APOE4* carrier and AD diagnosis (or Aβ status) as independent variables to account for the confounding effect of the *APOE4*-AD or *APOE4*-Aβ association: Model 1: Protein = β1 + β2 *APOE4*+ covariates + ɛ Model 2: Protein = β3 + β4 AD diagnosis (or Aβ) + covariates + ɛ Model 3: Protein = β5 + β6 *APOE4* + β7 AD diagnosis (or Aβ) + covariates + ɛ Covariates included age, sex, and mean overall protein level. Data contributor sites were included as an extra covariate for the GNPC cohort.

In order to investigate the protein profile of *APOE2*, we used the same 3 models on individuals containing only ε2 carriers (ε2ε2 and ε2ε3 carriers) compared to ε3ε3 carriers. In the 3 models, *APOE4* (ε3ε4+ε4ε4 vs. ε3ε3) was changed to *APOE2* (ε2ε2+ε2ε3 vs. ε3ε3), resulting in: Model 1: Protein = β1 + β2 *APOE2* + covariates + ɛ Model 3: Protein = β5 + β6 *APOE*2 + β7 AD diagnosis (or Aβ) + covariates + ɛ Covariates are the same as *APOE4* analysis.

To investigate early *APOE* effects, Model 1 was applied within the CU and Aβ-subgroups. To assess potential age-dependent variation in these associations, participants were stratified by median age into younger and older subgroups. Additionally, an *APOE*\*age interaction term was added to Model 1 within CU and Aβ-groups to statistically evaluate age-related modification of *APOE*-protein associations.

To investigate *APOE*-proteins associations change across different diagnostic or Aβ status groups, we also conducted separate analyses using Model 1 for the AD dementia (or Aβ+) groups. Similarly, an *APOE*\*AD dementia diagnosis (or *APOE*\*Aβ status) interaction term in Model 3 for the whole cohort was utilized to statistically assess whether the effects of *APOE* on proteins differ significantly between CU and AD individuals or between Aβ- and Aβ+ individuals.

To evaluate whether protein levels in CU and MCI *APOE4* carriers would alter their Aβ positivity rates, in BioFINDER-2 plasma SomaLogic cohort, we fit interaction models of the form: Aβ status = β1 + β2 *APOE4* + β3 protein + β4 (*APOE4*\*protein) + covariates + ɛ. Covariates included age, sex, and mean overall protein level, full statistical analysis available in Supplement Table 2. For proteins showing significant interaction with *APOE4*, we performed simple slopes analysis and calculated Johnson-Neyman intervals using the interactions R package (v1.2.0). This allowed us to identify the range of protein levels over which the effect of *APOE4* on Aβ status was statistically significant (α = 0.05), providing a more nuanced understanding of effect modification. In the simple slopes plot, the effect of *APOE4* on Aβ positivity was estimated at three protein expression levels: the mean, and ±1 standard deviation, with the slope representing the strength and direction of the association at each level.

All regression analyses were performed in R V.4.4.2. Graphs of individual protein levels grouped by *APOE* genotypes are the residuals after eliminating the effect of covariates on proteins using a regression model, statistical details can be found at Supplement Table 6. All statistical tests were two-sided, p values were adjusted using the Benjamini–Hochberg (FDR) correction method (for all multi-testing analysis), and significance was reported using FDR-corrected p values. Data visualizations were generated using the R package ggplot2 or the seaborn Python package V 0.13.2.

### Mediation analysis

To further explore the role of *APOE*-related proteins in AD clinical diagnosis or Aβ pathology, and to identify *APOE* (*APOE4* or *APOE2*) -related proteins affected by AD pathology or Aβ pathology, we tested them in different statistical mediation models. The mediation models were used to systematically classify *APOE*-related proteins into two protein groups: proteins that mediate the effects of *APOE* on AD clinical diagnosis or Aβ pathology (upstream mediation path: *APOE* => protein => AD diagnosis or Aβ status) and proteins that are affected by *APOE* through an AD-mediated pathway (downstream mediation path: *APOE* => AD diagnosis or Aβ status => protein). Our mediation study follows the Baron and Kenny’s Causal-Steps Test requirement, e.g. there is a significant correlation between the independent and dependent variables, i.e. only proteins associated with *APOE* in model 1 in the whole cohort were further tested for mediation models. Specifically, in the upstream mediation path, *APOE* was the exposure, AD diagnosis or Aβ status was the outcome, and *APOE*-associated proteins in model 1 (after FDR correction) were potential mediators. In the downstream mediation path, *APOE* was the exposure, *APOE*-associated proteins in model 1 (after FDR correction) were outcomes, and AD diagnosis or Aβ status was a potential mediator. Mediation analysis was performed using the “mediation” (V 4.5.0) R package where mediation proportions were estimated using causal mediation analysis with 95% confidence intervals based on 1,000 bootstrapping replicates. Direct effect, indirect effect and mediation proportion were extracted directly from model results and p values for these metrics were FDR corrected.

### BINN-enriched reactome pathway analysis

Biologically informed neural networks(BINNs)-enriched pathway analysis^65^ was used to globally annotate *APOE*-associated proteins that are altered already in CU or Aβ and its associated functionality. Specifically, based on the Reactome database (directed graph), the tool extracts the pathway hierarchy associated with the input protein and converts it into a neural network-like hierarchy (input *APOE* associated proteins → pathway layer → biological process layer → AD diagnosis or Aβ status). Finally, the Deep SHAP algorithm was used to calculate node importance and quantify the contribution of proteins/pathways to classification. The analysis is implemented in the binn Python package (V0.1.1). Proteins associated with *APOE4* or *APOE2* in both the whole cohort and CU (in GNPC, or Aβ- in BioFINDER-2 and ADNI) were conducted as input nodes separately. For proteins measured using multiple aptamers, we only kept the one with the smallest adjusted p value for *APOE* in model 1. In GNPC, the BINN model was trained to predict AD diagnosis. In BioFINDER-2 and ADNI, the model was trained to predict Aβ positivity in CU and MCI individuals to focus on early pathological changes; in ADNI, AD patients were included due to limited sample size. Data were split into training (80%) and validation (20%) sets, using a four-layer architecture trained over 100 epochs (500 epochs in ADNI due to smaller sample size).

### Subdivision of *APOE* associated proteins

To functionally stratify *APOE*-associated proteins, we categorized them into four groups based on their associations with AD diagnosis and mediation relationships:

1. Mediator proteins: Proteins showing significant indirect effects in the upstream mediation pathway (*APOE* => protein => AD diagnosis), but not in the reverse (*APOE* => AD diagnosis => protein), or for which the mediation proportion was stronger in the upstream than in the downstream direction (when both were significant after FDR correction). These proteins likely reflect early, genotype-driven molecular changes.
2. Pathology-mediated proteins: Proteins meeting the reverse pattern—i.e., stronger or exclusive mediation in the downstream pathway—suggesting they are regulated secondarily by AD.
3. *APOE*-specific proteins: Proteins significantly associated with *APOE* but not with AD diagnosis, potentially representing *APOE*-driven molecular changes independent of canonical AD diagnosis. Proteins falling into both this and mediation-related groups were classified according to their mediation status.
4. Non-specific proteins: Proteins associated with both *APOE* and AD, but not meeting mediation criteria in either direction.

For reference, we also defined a set of AD-specific proteins—those associated with AD diagnosis but not with *APOE* genotype—as putative markers of *APOE*-independent disease processes (Supplementary Table 2). In analyses focused on Aβ pathology, classification of *APOE*-associated proteins into the four functional categories was performed analogously, based on their associations with Aβ status and respective mediation pathways.

### Linear Discriminant Analysis

To assess whether the identified functional categories of *APOE*-associated proteins correspond to distinct molecular profiles, we evaluated their separability using Linear Discriminant Analysis (LDA) in the GNPC cohort. Specifically, an LDA model was trained using the five predefined categories—mediator proteins, pathology-mediated proteins, *APOE*-specific proteins, non-specific proteins, and AD-specific proteins—as class labels. Feature vectors were constructed from protein expression residuals after adjusting for age, sex, mean protein level, and data collection site, to isolate *APOE* and *AD* related variance. The model, implemented using the scikit-learn Python package (v1.3.1), was trained on this labeled subset and used to derive two linear discriminants (LD1 and LD2) capturing maximal between-class variance for visualization. After training, the entire proteome (7,285 proteins) was projected into the learned discriminant space to evaluate whether proteins in different functional categories formed separable clusters. Cluster separation and within-group compactness were qualitatively assessed in the resulting 2D projection.

### Cell type and functional enrichment analysis

To further investigate the potential cellular and function origins of *APOE*-associated proteomic changes, we performed cell-type enrichment analysis for genes corresponding to proteins in each of the four functional categories defined above.

For cell-type enrichment analysis, we used two independent single-cell RNA sequencing (scRNA-seq) datasets: 1) 2.3 million single-cell transcriptomes from the aged human prefrontal cortex, collected from 427 participants in the Religious Orders Study (ROS) and Rush Memory and Aging Project (MAP)^66^, spanning neuronal, glial, and vascular populations; and 2) the Human Brain Vascular Atlas^67^, focused specifically on vascular and perivascular cell types, including 143,793 single-cell transcriptomes from the hippocampus and cortex of eight post-mortem human brains.

For both datasets, we obtained the Seurat objects and processed them using the R package Seurat (version 4.3.0)^68^. We applied the AverageExpression function to compute the average gene expression levels for each cell type. Subsequently, we calculated the proportion of gene expression distributed across major neuronal and non-neuronal cell populations.

To assess whether specific protein sets showed preferential expression in particular cell types, we computed the mean expression levels of the significant protein within each protein group across all cell types. To evaluate whether the observed enrichment exceeded what would be expected by chance, we generated 10,000 random gene lists sampled from the SomaLogic background set, each matched in size to the significant list. Using these random sets, we constructed a probability distribution of average expression levels across cell types. This approach allowed us to estimate the probability that the observed enrichment was greater than random expectation, accounting for background expression variability. Cell types were considered significantly enriched if the false discovery rate (pFDR), corrected using the Benjamini–Hochberg method, was below 0.05.

Functional enrichment analyses were performed using Gene Ontology (GO) Biological Process (BP) terms. We conducted an overrepresentation analysis with the enrichGO() function from the clusterProfiler package^69^ (v4.6.2) and org.Hs.eg.db annotation^70^ database (v3.16.0) in R. As individual significant protein groups did not yield enrichment at a false discovery rate (pFDR) threshold of 0.05, and to maximize biological interpretability by capturing the broader functional context of the identified proteins, we conducted functional enrichment analysis on two complementary protein sets: (1) Set 1, consisting of proteins significantly upregulated or downregulated by *APOE* within each protein group; and (2) Set 2, consisting of proteins significantly upregulated or downregulated by *APOE* within each protein group together with their first-degree protein-protein interaction (PPI) partners. First-degree PPI partners were identified using the InWeb database^71^ of measured and inferred physical interactions, restricting analyses to high-confidence (’gold-standard’) interactions. The background set consisted of all proteins quantified by the SomaLogic platform combined with the first-degree protein-protein interaction partners of the significant proteins identified in our study. To increase the robustness of the results and minimize the likelihood of spurious findings, we focused on biological processes that were enriched in both Set 1 and Set 2. We selected nominally significant terms (nominal p<0.05) from Set 1 and FDR-corrected significant terms (pFDR<0.05) from Set 2, identified overlapping biological processes between the two sets, reduced redundancy among GO terms, and manually summarized the top processes into major biological processes. All significant overlapping biological processes are reported in Supplementary Table 4.

Finally, protein-protein interaction networks were constructed by querying the STRING database separately for the full list of significant proteins associated with *APOE4* or *APOE2*, using a minimum combined score threshold of 0.7 to ensure high-confidence interactions.

### Sensitivity analyses

While primary analyses of the GNPC and BioFINDER-2 cohorts were conducted independently—with GNPC focused on clinical AD diagnosis and BioFINDER-2 on Aβ pathology—we noted partial sample overlap between the two cohorts. To ensure fair cross-cohort comparisons and avoid artificial inflation of concordance, we performed a sensitivity analysis in which overlapping individuals were excluded from the GNPC dataset prior to comparison with BioFINDER-2 results. Association and mediation analyses were then re-conducted in the reduced GNPC sample, and the results were compared to those from BioFINDER-2 to assess the robustness and consistency of findings across non-overlapping populations (Supplement Fig. 8A, B; Supplement Table 2).

As for the BioFINDER-2 cohort, we utilized available SNP data and performed principal component analysis (PCA) using PLINK v2.0 to account for potential population stratification. The top 10 principal components (PCs) were calculated based on genome-wide SNP data (with LD filtering, MAF threshold and other pre-processing steps). Among them, the first five PCs (PC1–PC5), which captured the major axes of genetic variation, were included as covariates for BioFINDER-2 data in a separate analysis to correct for population structure. Results for sensitive analysis are available in Supplement text and figures.

### Regional gene expression analysis

To further investigate the relation between *APOE* and related proteins, we performed region-wise association between gene expression of the proteins related to *APOE4* or *APOE2*, and regional gene expression of *APOE* or regional Aβ-PET, based on the Schaefer atlas with 200 parcels. Gene expression was generated using the regional microarray expression data obtained from 6 post-mortem brains provided by the Allen Human Brain Atlas^72^ (https://human.brain-map.org). Data were processed with the abagen toolbox version 0.1.3^72–74^ (https://abagen.readthedocs.io/en/stable/index.html), a Python-based package to access and work with the Allen Brain data microarray expression data. To ensure analyses were not driven by statistical autocorrelations, we performed null modeling with BrainSMASH (https://brainsmash.readthedocs.io/en/latest/index.html), a Python-based package for statistical testing of spatially autocorrelated brain maps^75^. BrainSMASH simulates surrogate brain maps with spatial autocorrelation that matches the spatial autocorrelation of the original brain map of interest, in this case the *APOE* or the Aβ-PET map. For both software, we used the default parameters and followed the same steps as described previously^22^. Non-parametric *p*-values were computed, corresponding to the frequency that correlation with the surrogate maps exceeded the observed correlation with the original gene expression map. *APOE4*- or *APOE2*-related proteins were considered to have gene expression profiles significantly related to *APOE* when BrainSMASH-corrected p-values after FDR correction was smaller than 0.05.

### Longitudinal analysis

To explore how proteomic trajectories change over time in *APOE4* or *APOE2* carriers. We examined the interaction between time and *APOE* status in the PPMI cohort (CSF OLINK), adjusting for age, sex, mean overall protein levels, and the diagnostic group using linear mixed-effects (LME) models. Separate models were fitted to assess the interactive effects of time with *APOE4* (comparing ε4 carriers to ε3ε3 carriers) or *APOE2* (comparing ε2 carriers to ε3ε3 carriers) excluding ε2ε4 carriers. These analyses were conducted across all 826 proteins, full results are available in Supplement Table 5. In the main text, we focused on a subset of proteins associated with *APOE* in GNPC or in BioFINDER-2 CSF OLINK baseline analyses.

## Data availability

GNPC (https://www.neuroproteome.org/) will be open-access upon the publication of this study. ADNI, UKB and PPMI data used in this manuscript are publicly available from the ADNI database (<u>adni.loni.usc.edu</u>), UKB database (https://www.ukbiobank.ac.uk/) and PPMI database (https://www.ppmi-info.org/) on registration and compliance with the data use agreement. GWAS summary data was publicly available in https://www.ebi.ac.uk/gwas/.

Pseudonymized data from the BioFINDER study (Principal Investigator: O.H.) are available to qualified academic researchers upon request, specifically for the purpose of replicating the analyses reported in this study. In accordance with the EU General Data Protection Regulation (GDPR), access to these data requires a data transfer agreement with Skåne University Hospital (Region Skåne), which outlines provisions for secure data handling, storage, and usage. All proposed analyses must comply with the ethical approvals granted by the Swedish Ethical Review Authority. This controlled access ensures the confidentiality of study participants, who did not consent to unrestricted public data sharing, and ensures that data use remains consistent with the scope of the original ethical approvals.

## Code Availability

Essential code for the manuscript can be found at https://github.com/Lina0125/APOE_proteomics.

## Data Availability

GNPC (https://www.neuroproteome.org/) will be open-access upon the publication of this study. ADNI, UKB and PPMI data used in this manuscript are publicly available from the ADNI database (adni.loni.usc.edu), UKB database (https://www.ukbiobank.ac.uk/) and PPMI database (https://www.ppmi-info.org/) on registration and compliance with the data use agreement. GWAS summary data was publicly available in https://www.ebi.ac.uk/gwas/. Pseudonymized data from the BioFINDER study (Principal Investigator: O.H.) are available to qualified academic researchers upon request, specifically for the purpose of replicating the analyses reported in this study. In accordance with the EU General Data Protection Regulation (GDPR), access to these data requires a data transfer agreement with Skane University Hospital (Region Skane), which outlines provisions for secure data handling, storage, and usage. All proposed analyses must comply with the ethical approvals granted by the Swedish Ethical Review Authority. This controlled access ensures the confidentiality of study participants, who did not consent to unrestricted public data sharing, and ensures that data use remains consistent with the scope of the original ethical approvals.

https://github.com/Lina0125/APOE_proteomics

## Acknowledgments

This research has been conducted using the UK Biobank Resource under Application Number 105777. The BioFINDER study was supported by the National Institute of Aging (R01AG083740), European Research Council (ADG-101096455), Alzheimer’s Association (ZEN24-1069572, SG-23-1061717, ALZSI-26-1523522), GHR Foundation, Swedish Research Council (2022-00775, 2021-02219, 2019-2024), ERA PerMed (ERAPERMED2021-184), Knut and Alice Wallenberg foundation (2022-0231), Strategic Research Area MultiPark (Multidisciplinary Research in Parkinson’s disease) at Lund University, Swedish Alzheimer Foundation (AF-980907, AF-994229, AF-1011949, AF-1011799), Swedish Brain Foundation (FO2021-0293, FO2023-0163, FO2024-0284, FO2025-0055), WASP and DDLS Joint call for research projects (WASP/DDLS22-066), Parkinson foundation of Sweden (1412/22), Cure Alzheimer’s fund, Rönström Family Foundation, Berg Family, Bundy Academy, Konung Gustaf V:s och Drottning Victorias Frimurarestiftelse, Skåne University Hospital Foundation (2020-O000028), Regionalt Forskningsstöd (2022-1259), Michael J Fox Foundation (MJFF-025507), Lilly Research Award Program and Swedish federal government under the ALF agreement (2022-Projekt0080, 2022-Projekt0107). Author JWV was supported by the SciLifeLab & Wallenberg Data Driven Life Science Program (grant: KAW 2020.0239) and the Swedish Research Council (2024-03642). The precursor of 18F-flutemetamol was sponsored by GE Healthcare. The precursor of 18F-RO948 was provided by Roche. The funding sources had no role in the design and conduct of the study; in the collection, analysis, interpretation of the data; or in the preparation, review, or approval of the manuscript.

## Ethics disclosures

### Conflicts of interest

NMC has received speaker/consultancy fees from Biogen, Eli Lilly, Owkin and Merck. OH is an employee of Lund University and Eli Lilly. SP has acquired research support (for the institution) from Avid and ki elements through ADDF. In the past 2 years, he has received consultancy/speaker fees from Bioartic, Biogen, Eisai, Eli Lilly, Novo Nordisk, and Roche. BM and AV are employees of Abbvie. JWV has received consultancy fees from Manifest Technologies.

## Notes

### Author Declarations

Ethical approval for the Swedish BioFINDER-2 cohort (https://clinicaltrials.gov/study/NCT03174938) was obtained from the Regional Ethical Committee in Lund, Sweden.

### Summary of Updates

Section on abstract updated to clarify usage and findings of each cohort; Acknowledgement updated; Supplemental figures and texts were updated.

