## Supplement Text and Figures for "Proteomic signatures of the *APOE ε4* and *APOE ε2* genetic variants and Alzheimer’s disease"

### The GNPC discovery cohort

#### *APOE4*-associated proteins

**Supplement Fig .1 : *APOE4*-associated proteins in GNPC (plasma Somalogic)**

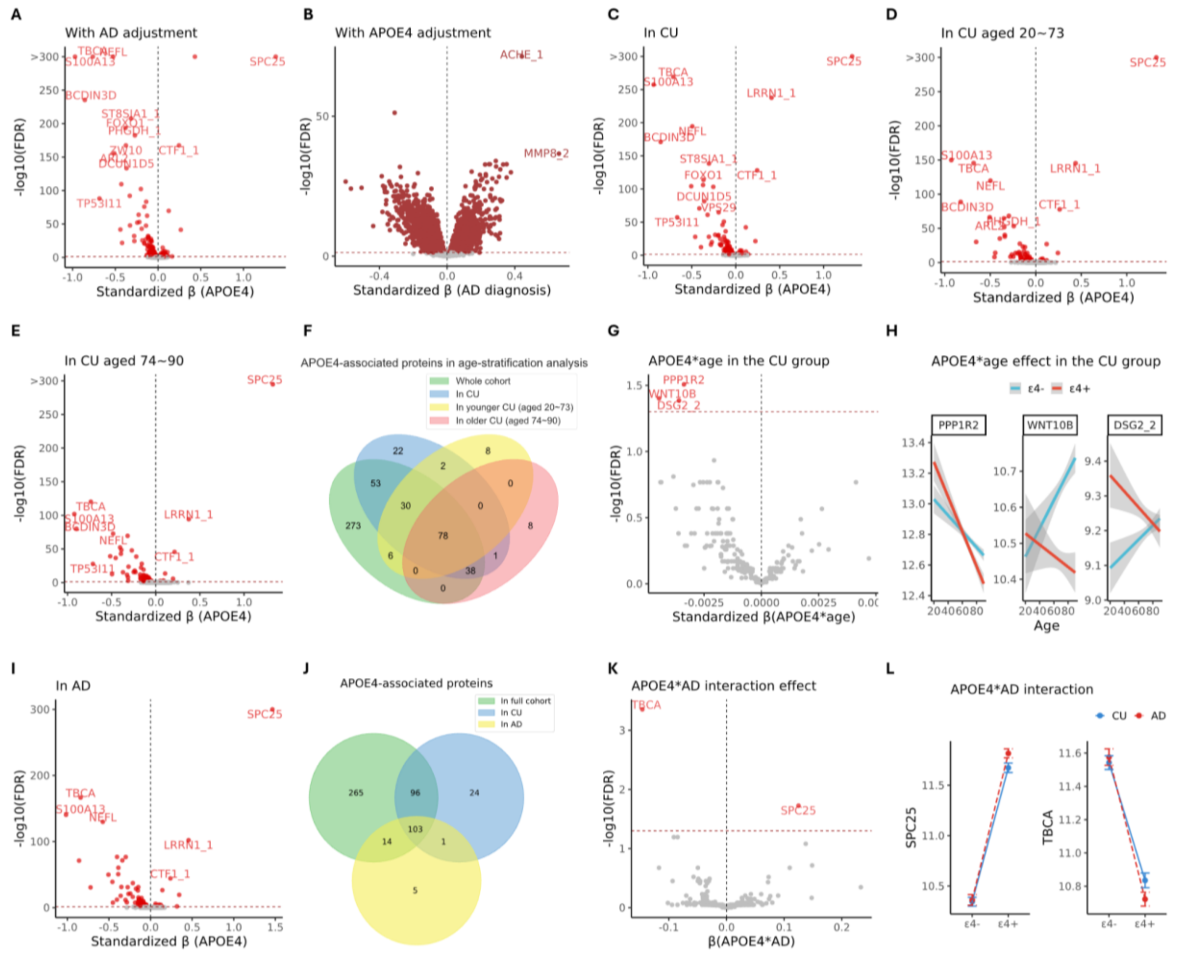

**A**. Volcano plot for proteins associated with *APOE4* with adjusting for AD diagnosis. At y-axis, -log10(FDR) above 300 was set to 300 for a better visualization. For all volcano plots, red represents significant association after FDR correction. **B**. Volcano plot shows proteins associated with AD diagnosis with adjusting for *APOE4*. **C**. Volcano plots show proteins associated with *APOE4* in CU. **D**. Volcano plots show proteins associated with *APOE4* in younger CU stratified using median age of CU group. **E**. Volcano plots show proteins associated with *APOE4* in older CU stratified using median age of CU group. **F**. Venn plot shows intersection of *APOE4* associated proteins in whole cohort, in CU and stratified age groups. **G**. Volcano plot shows *APOE4**age effect on proteins in CU. **H**. Interaction effect of *APOE4* status and age on protein levels in the CU group. Protein levels of PPP1R2, WNT10B and DSG_2 ("_2" represent one aptamer for DSG) are plotted against age, with separate regression lines for individuals carrying the ε4 allele (ε4+, red) and those without it (ε4-, blue). Shaded areas represent 95% confidence intervals. **I**. Volcano plot shows proteins associated with *APOE4* in the AD dementia group. **J**. Venn plot shows the number of proteins associated with *APOE4* in the whole cohort, in CU and in the AD dementia group. **K**. Volcano plot shows *APOE4**AD diagnosis interactive effect on proteins. **L**. Interaction plots show how *APOE4*'s effect on protein level changes in AD diagnosis groups compared to the CU group.

#### *APOE2*-associated proteins

**Supplement Fig . 2 : *APOE2*-associated proteins in GNPC (plasma Somalogic)**

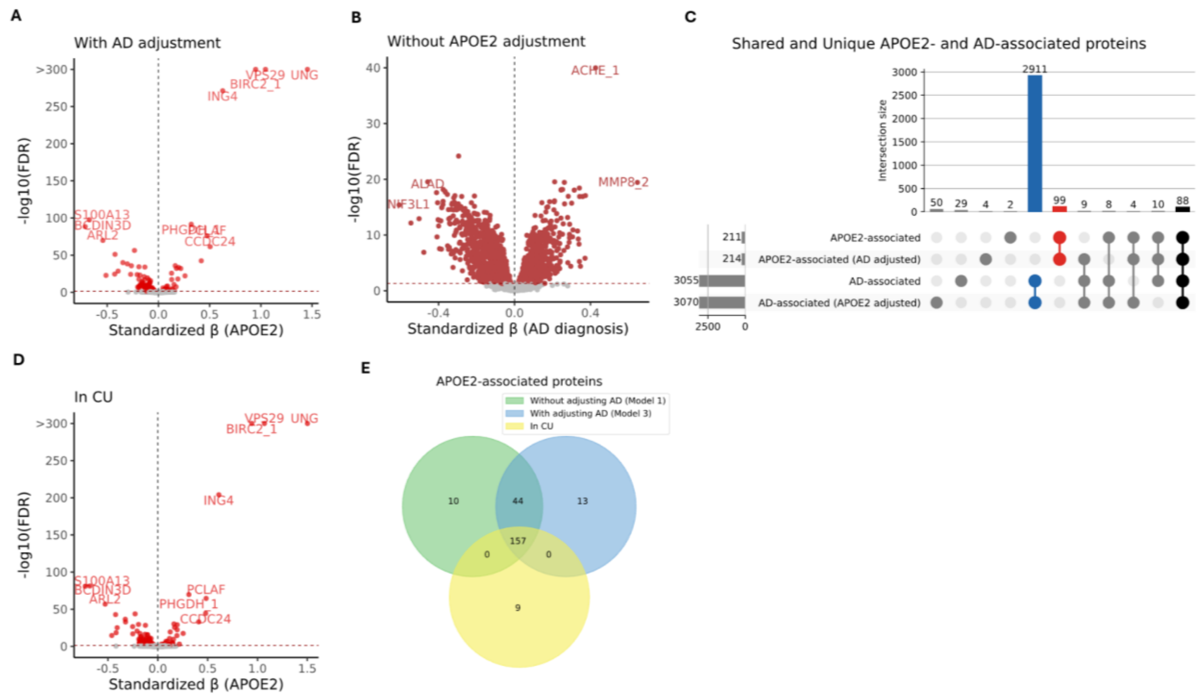

**A**. Volcano plot shows proteins associated with *APOE2* with AD adjustment. For volcano plots, red dots represent a significant association. **B**. Volcano plot shows proteins associated with AD without adjusting *APOE4*. **C**. The UpSet plot shows the number of proteins associated with *APOE2* or AD with or without adjusting each other. Blue indicates the number of proteins specific to AD while red indicates the number of proteins specific to *APOE2*. **D**. Volcano plot shows proteins associated with *APOE2* in the CU group. For both plots, red represents a significant association after FDR correction. **E**. Venn plot shows the number of proteins associated with *APOE2* in model 1 (without adjusting AD), model 3 (with adjusting AD) and in CU.

### Overview of proteins in each validation dataset

#### Shared proteins in each dataset

6,358 unique proteins were included for the GNPC and BioFINDER-2 plasma SomaLogic cohort, 800 proteins were measured using multiple aptamers, resulting in 7,285 aptamers in total. Similarly, 6,135 unique proteins were included in ADNI CSF SomaLogic cohort, 754 proteins were measured using multiple aptamers, resulting in 7,001 aptamers in total. 1,382 unique proteins were included in the BioFINDER-2 CSF OLINK cohort, 3 proteins were measured using 4 separate panels respectively, resulting in 1,391 OLINK NPX measurements. 1,319 proteins were included and no protein was measured using multiple panels for the UKBB cohort.

**Supplement Fig . 3: Shared proteins in each dataset**

**A**

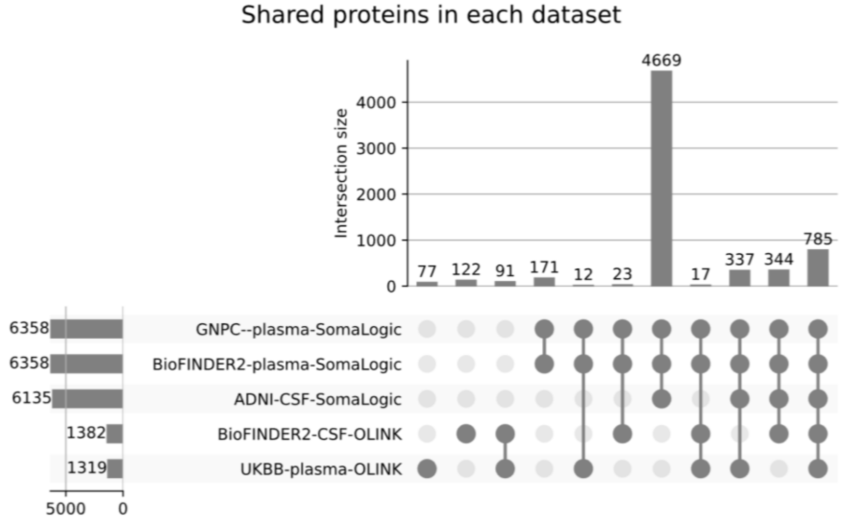

**A**. UpSet plot shows the number of unique proteins included in each dataset.

### BioFINDER-2 plasma SomaLogic cohort

#### Validation of early *APOE*-linked pathways

**Supplement Fig . 4: Validation of early *APOE*-linked pathways (BioFINDER-2 plasma SomaLogic)**

**A**

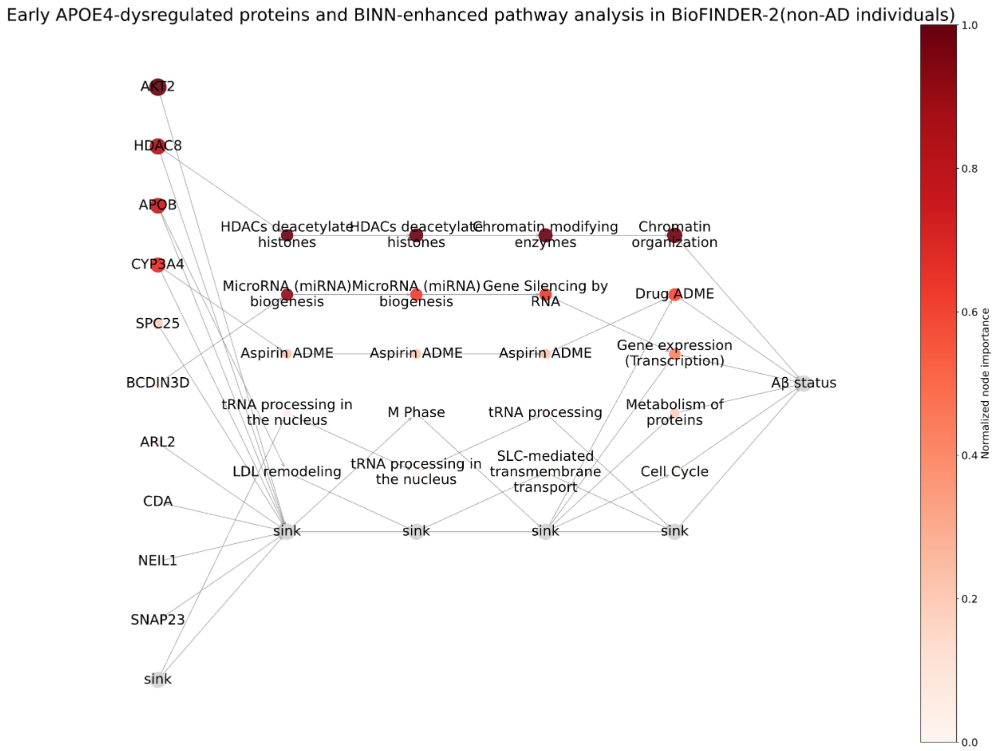

**B**

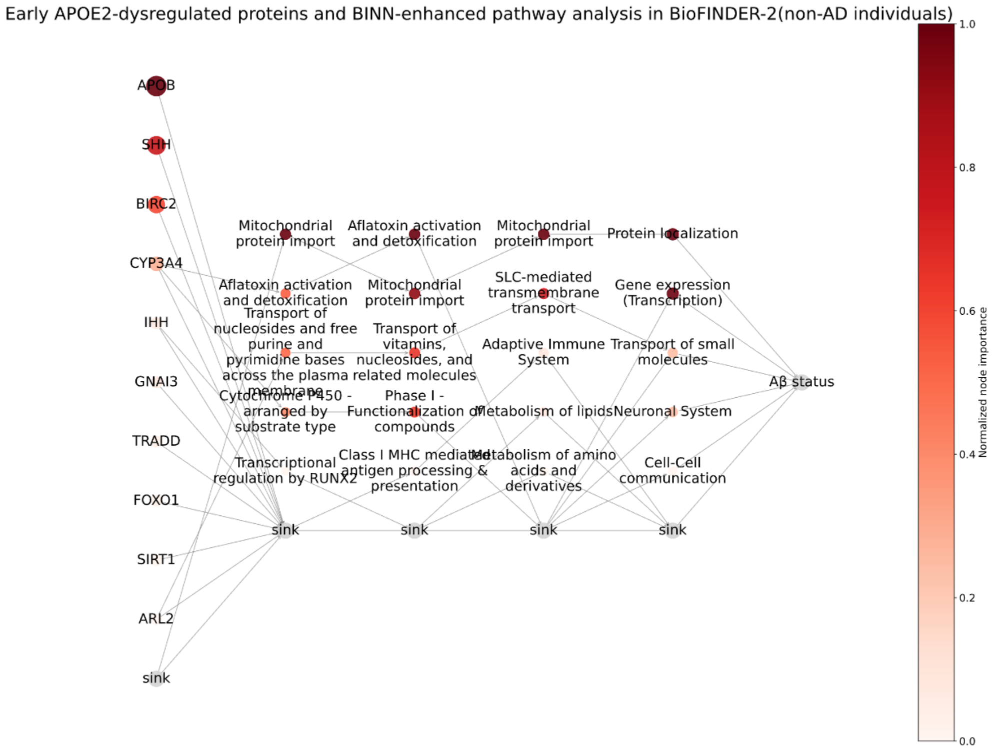

BINN-enriched reactome pathway analysis for proteins associated with *APOE4* (**A**) or *APOE2* (**B**) in both the whole cohort and in the Aβ- group in the BioFINDER-2 plasma SomaLogic cohort. The trained model showed good performance in predicting Aβ status in CU and MCI individuals: 0.7 training and testing accuracy when using *APOE4* associated proteins as input nodes (**A**), 0.66 training and 0.64 testing accuracy when using *APOE2* associated proteins as input nodes (**B**). The plot shows the most important proteins and associated pathways in the deep learning models predicting Aβ status in CU and MCI participants. The darker the dot, the more important the protein and the pathway in the deep learning model predicting Aβ status. More features are hidden in the sink for a better visualization.

#### Sensitive analysis: adjusting for population stratification

In order to eliminate the potential population confounding, we calculated the top 10 principal components (PCs) based on genome-wide SNP data and adjusted PC1-5 as covariates as sensitive analysis in the BioFINDER-2 plasma SomaLogic dataset. 163 proteins associated with *APOE4* after adjusting PC1-5, including key proteins e.g. SPC25, etc., 150 were consistent with the main analysis (Supplement Table 2). 69 associated with *APOE4* in Aβ- after adjusting PC1-5, including key proteins e.g. SPC25 etc., 64 were consistent with the main analysis. 7 mediators (SPC25, TBCA, S100A13, CTF1, BCDIN3D, CDA and DCUN1D5) in *APOE4* => protein => Aβ pathway were replicated from the main analysis, although estimation of mediation proportion is not significant, indicating an unstable estimate (Supplement Fig. 5A). 8 proteins mediating *APOE4*'s effect on AD diagnosis after adjusting genetics PC1-5, with 5 e.g. TBCA, S100A13 and PHGDH replicated from the main analysis (Supplement Fig. 4B).

No proteins found to mediate the effect of *APOE2* on Aβ or AD diagnosis after adjusting for top 5 genetic PCs. Full statistical analysis can be found at Supplement Table 2.

**Supplement Fig . 5: Sensitive analysis in BioFINDER-2 plasma SomaLogic**

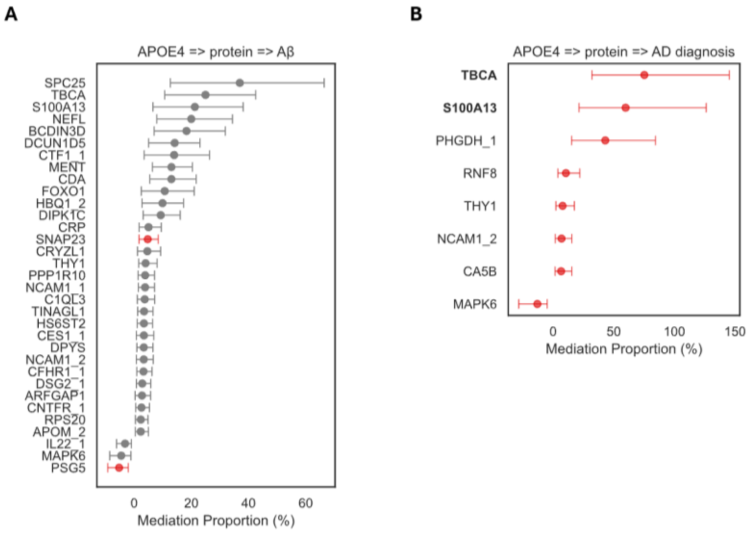

The dot plot with error bars shows the mediation proportions and confidence intervals of proteins mediating the effects of *APOE4* on Aβ (**A**) and on AD clinical diagnosis (**B**) after adjusting for population stratification (top 5 genetic PCs). Proteins whose indirect effects in protein mediation pathways are significant alone or mediation proportions of protein-mediation pathways are greater than those of Aβ (or AD) -mediated pathways (when indirect effects of both mediation paths are significant) are defined as mediators and are shown. The x-axis represents the percentage of mediation proportions. The dots represent the estimated mediation proportions of each protein while red indicates a significant estimation and grey indicates a non-significant estimation. The horizontal lines represent the 95% confidence intervals of these estimates. Bold indicates that the direct effect of *APOE4* is not significant, resulting in a total mediation effect of this protein.

### ADNI CSF SomaLogic cohort

#### Validation of early *APOE*-linked pathways in CSF

**Supplement Fig . 6: Validation of early *APOE*-linked pathways (ADNI CSF SomaLogic)**

**A**

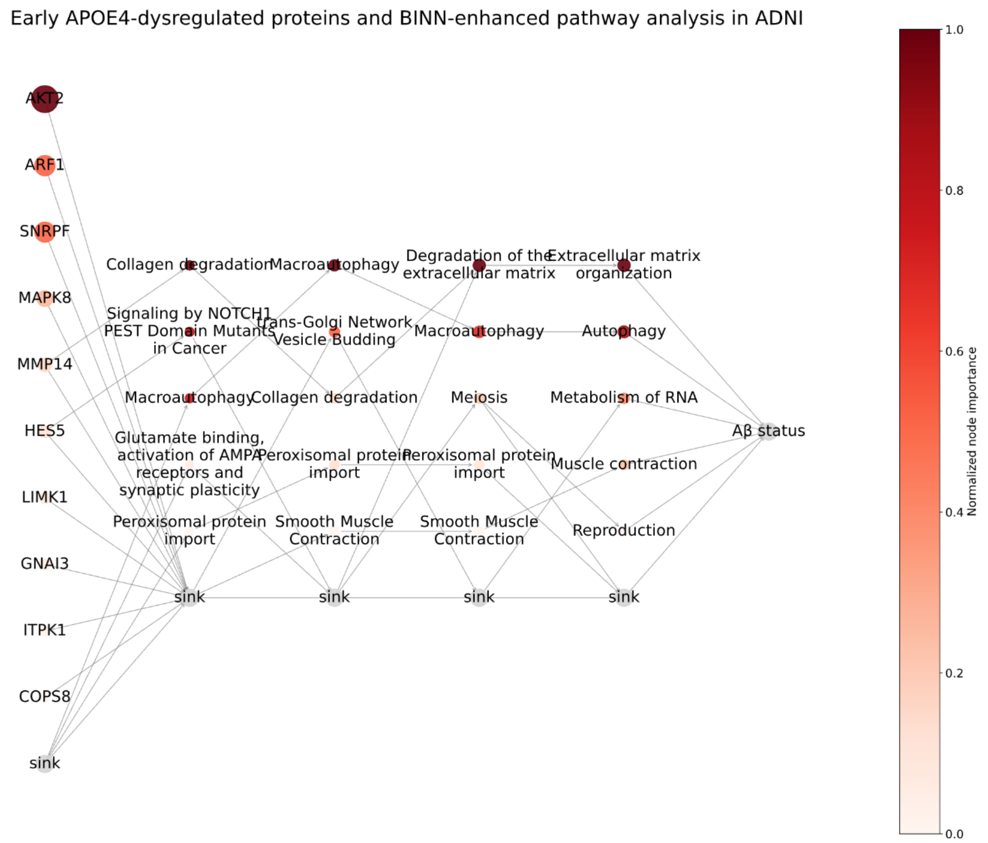

**B**

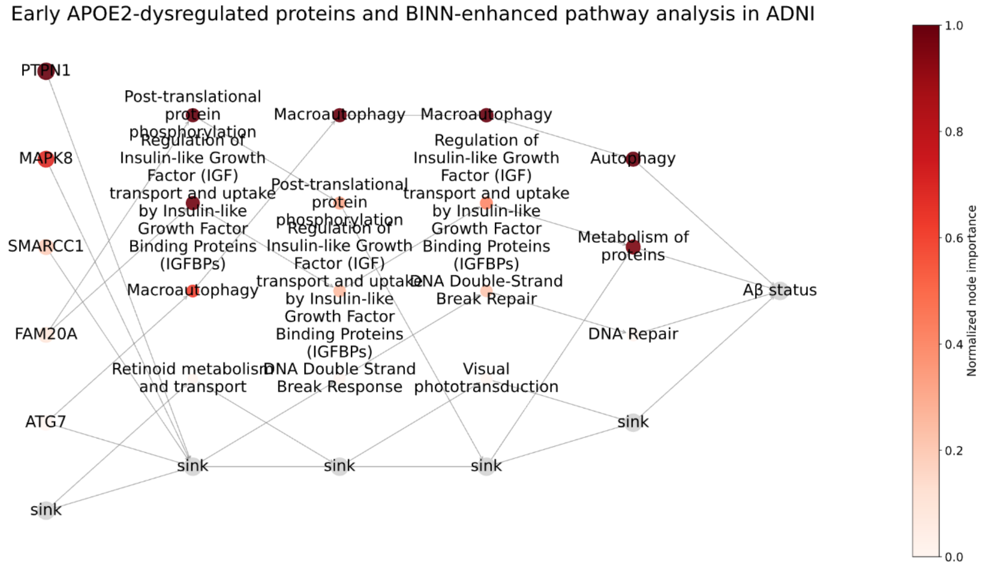

BINN-enriched reactome pathway analysis for proteins associated with *APOE4* (**A**) or *APOE2* (**B**) in both the whole cohort and in Aβ- individuals. The plot shows the most important proteins and associated pathways in the deep learning models predicting Aβ status in CU, MCI and AD (due to the small sample size of Aβ+ individuals, the AD group was included) participants. The darker the dot, the more important the protein and the pathway in the deep learning model predicting AD dementia diagnosis. More features are hidden in the sink for a better visualization.

### UKBB plasma OLINK cohort

#### Validation of *APOE*-plasma proteomics signatures in OLINK

**Supplement Fig . 7: Validation of APOE-plasma proteomics signatures in OLINK (UKBB)**

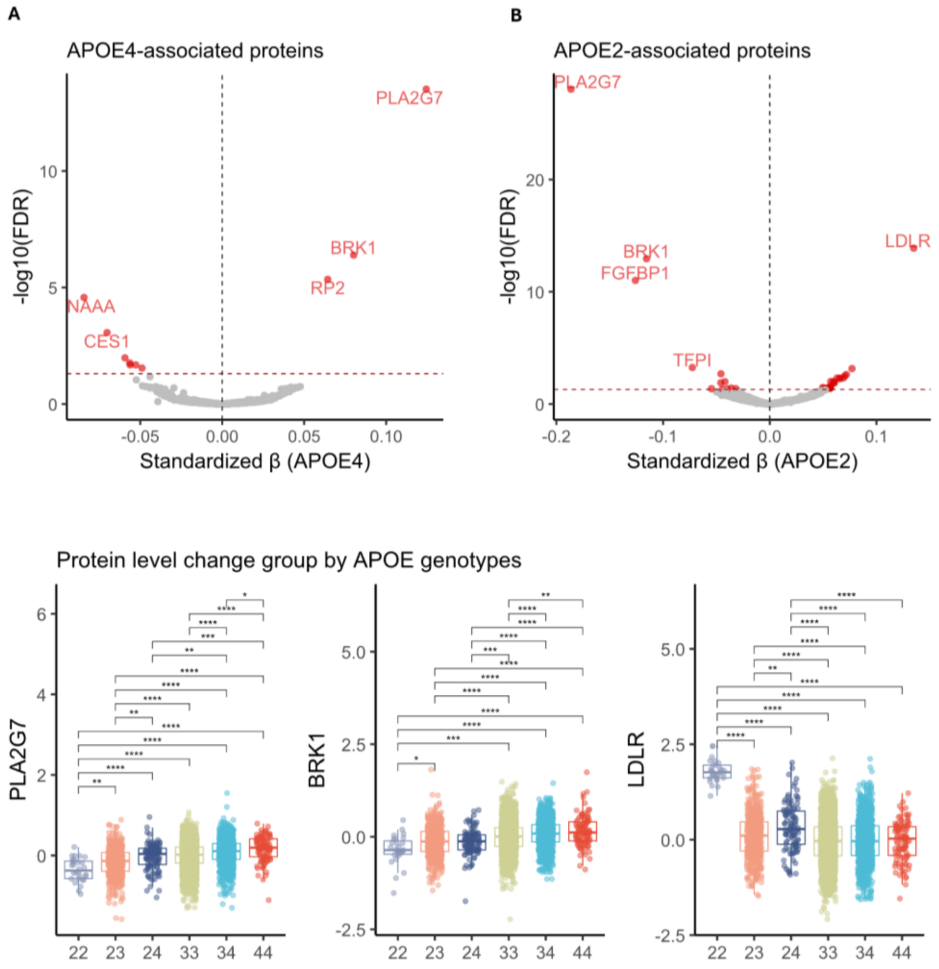

**A**. Volcano plot shows proteins associated with *APOE4* without AD or Aβ adjustment (unavailable for testing). For volcano plots, red indicates a significant association after FDR corrections. **B**. Volcano plot shows proteins associated with *APOE2* without AD or Aβ adjustment (unavailable for testing) **C**. Boxplots shows protein level change groups by detailed *APOE* genotypes for key proteins associated with *APOE*.

### Longitudinal *APOE*-protein associations in PPMI

Longitudinal *APOE*-protein associations were investigated using the PPMI CSF OLINK proteomics dataset. A total of 253 individuals were included after excluding 7 ε2ε4 carriers. Among them, 65 were *APOE4* carriers and 30 were *APOE2* carriers. The cohort comprised 74 Parkinson's disease (PD) patients, 56 prodromal individuals, and 130 healthy controls; 205 were CU and 18 had MCI. We included non-AD pathology individuals due to a small sample size.

A total of 826 proteins passed quality control and were analyzed using linear mixed-effects (LME) models to test for *APOE4**time and *APOE2**time interaction effects, adjusting for age, sex, mean protein expression, and diagnostic group. We focused on proteins previously found to be associated with *APOE* in previous analysis; results for all proteins are provided in Supplement Table 5.

Of the 72 GNPC *APOE4*-associated proteins available in the PPMI dataset, LGALS9 and PRSS8—both downstream of AD in ε4 and ε3ε3 carriers in GNPC—showed nominally significant *APOE4**time interactions, with levels increasing faster over time in *APOE4* carriers. Among the 12 *APOE2*-associated proteins, only NEFL showed a nominal *APOE2**time interaction, with a faster increased level over time in *APOE2* carriers.

To address potential platform and tissue differences, we further assessed 25 *APOE4* and 1 *APOE2* associated protein (LDLR) in the BioFINDER-2 CSF OLINK dataset. Only SIGLEC1 showed a nominally significant *APOE4**time interaction, with SIGLEC1 levels in CSF increasing faster over time in *APOE4* carriers.

However, the limited longitudinal changes observed may reflect platform- or matrix-specific differences, or limited statistical power, rather than true absence of dynamic changes, highlighting the need for larger-scale longitudinal cohorts to better capture the temporal trajectory of *APOE*-mediated proteomic shifts.

### Heterogeneity across platforms and tissues in *APOE* proteomic signatures

To investigate the cross-cohort and cross-platform heterogeneity of proteomic signatures, we used adjusted $R^2$ and effect size of *APOE4* as the matrices to evaluate the generalizability of model fitting and the effect of *APOE4* on proteins across all dataset, specifically, we calculated the Spearman correlation of adjusted $R^2$ values and effect size of *APOE4* (from Model 1, as Model 1 is the model that generally used in all dataset) for each protein across datasets (for proteins that could be matched). Noting that for the results used comparing different datasets, we conducted separate analysis excluding participants from BioFINDER-2 cohort in GNPC to eliminate possible bias, the results can be found at Supplement Table 8.

For model fitting, The GNPC plasma SomaLogic (overlapping subjects with the BioFINDER-2 cohort were excluded) and BioFINDER-2 plasma SomaLogic cohorts showed the strongest agreement among 6,358 shared proteins (r = 0.72, Supplement Fig. 8A). Moderate correlations were also observed between BioFINDER-2 plasma SomaLogic and UKBB plasma OLINK (1,151 proteins, r = 0.4), GNPC plasma SomaLogic and UKBB plasma OLINK (1,151 proteins, r = 0.38). A significant correlation was found between the two CSF cross-platform datasets, ADNI CSF SomaLogic and BioFINDER-2 CSF OLINK (1,129 proteins, r = 0.37). The same platform with different tissues including plasma (GNPC, BioFINDER-2) vs. CSF (ADNI) in SomaLogic and plasma (UKBB) vs. CSF (BioFINDER-2) in OLINK both showed significant but limited correlation (r = 0.14 ~ 0.17). Correlations between dataset using different tissues and different proteomics platforms remained low or showed no correlations even for the same cohort, like CSF OLINK vs. plasma SomaLogic in the BioFINDER-2 cohort (Supplement Fig. 8A).

However, when it comes to the correlation of the effect size of *APOE4* on shared proteins, GNPC and BioFINDER-2 plasma SomaLogic had only medium correlation (r = 0.33), ADNI CSF SomaLogic had smaller correlation with BioFINDER-2 CSF OLINK (r = 0.26) and limited correlation with BioFINDER-2 plasma SomaLogic (r = 0.14) and GNPC plasma SomaLogic (r = 0.14), reflecting large differences (Supplement Fig. 8B).

To further evaluate measurement concordance, we further analyzed 1,169 shared proteins in 1,349 individuals from BioFINDER-2 who had both plasma SomaLogic and CSF OLINK data. After adjusting platform-specific mean levels, the overall cross-platform and cross-tissue correlation remained low, although CSF OLINK showed relatively higher autocorrelation than plasma SomaLogic measurements (Supplement Fig. 8C), possibly indicating a higher protein variance in plasma. Protein-wise analysis revealed that for 88.5% of proteins, the absolute correlation between CSF (OLINK) and plasma (SomaLogic) levels was below 0.3 (Supplement Fig. 8D). Only a few proteins, such as PNLIPRP2 and CHFR4, showed strong correlations on the 2 measurements from plasma SomaLogic and CSF OLINK (Supplementary Fig. 8E).

We observed different results for individual proteins from each dataset; a representative example is NEFL. Specifically, plasma NEFL ANML(SomaLogic) level is mediating *APOE4*'s effect on AD diagnosis (GNPC) and Aβ status (after adjusting for population stratification in the BioFINDER-2 cohort) and was significantly downregulated in *APOE4* carriers and *APOE2* carriers (even in CU and Aβ-), AD and Aβ+ individuals (although not in *APOE2* analysis only contains ε2 carriers and ε3ε3 carriers). While CSF NEFL (both OLINK NPX level and SomaLogic ANML level) was only associated with Aβ status (upregulated in Aβ+) but not with *APOE4* or *APOE2.* We next verified the association between *APOE* genotype and plasma neurofilament light (NfL) level measured using the Quanterix Simoa® NfL assay and CSF NfL level measured using Roche Elecsys NeuroToolKit (NTK) assay. After eliminating the effect of age, sex, mean protein level (not for Simoa and NTK), and Aβ status, only NEFL measured using plasma SomaLogic were associated with *APOE* genotypes, no significant association between *APOE* with either CSF NEFL measured using SomaLogic or OLINK, CSF NfL measured using Roche Elecsys NeuroToolKit (NTK) assay or plasma NfL measured using the Quanterix Simoa® NfL assay observed (Supplement Fig. 8F).

We next calculated the correlations between different NEFL measurements in the BioFINDER-2 cohort. Plasma NfL (Simoa) and CSF NfL (NTK) showed the highest positive correlations (Spearman r=0.77, FDR<0.05). No significant associations observed between other NEFL measurements (Supplement Fig. 8G).

Despite robust findings of mediation results, no protein found to be consistently associated with *APOE4* across all 5 datasets (Supplement Fig. 8H). Similarly, no protein was consistently associated with *APOE2* in all 5 datasets. (Supplement Fig. 8I).

**Supplement Fig. 8：Commence and heterozygosity of *APOE* proteomics across platforms and tissues**

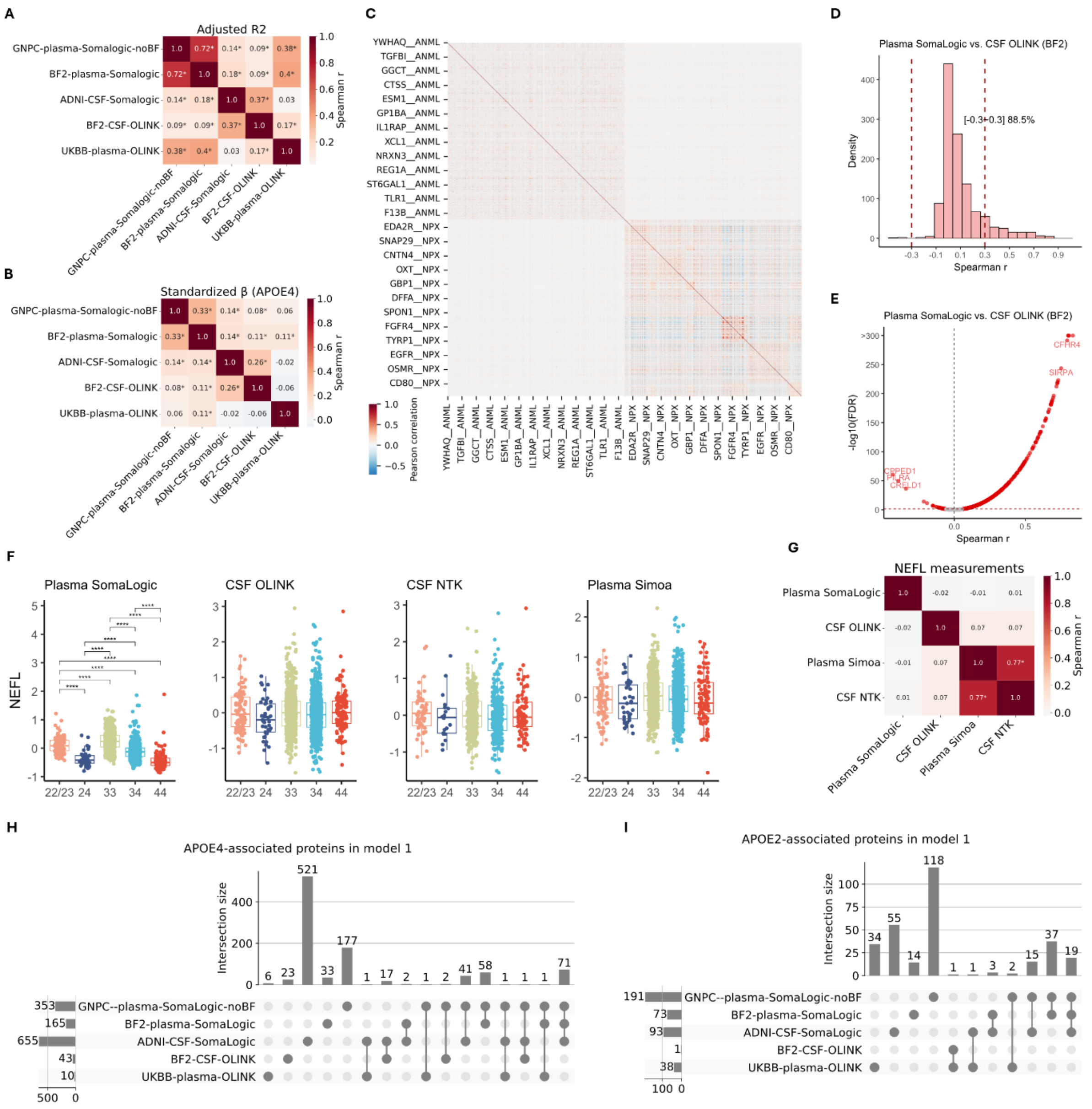

**A**. Heatmap shows the spearman correlation of adjusted R-square of model 1 (*APOE4*) on shared proteins in 2 datasets. **B**. Heatmap shows the spearman correlation of effect size of *APOE4* in model 1 on shared proteins in 2 datasets. **C**. Cluster plot shows the autocorrelation and correlation across plasma SomaLogic proteins (ANML) and CSF OLINK (NPX) for 1,168 proteins in 1,388 subjects in the BioFINDER-2 cohort. **D**. Histogram shows the distribution of spearman r of protein level measured using plasma SomaLogic and CSF OLINK in BioFINDER-2 cohort. **E**. Volcano plot shows detailed spearman R vs. p value for proteins. **F**. Boxplots show NEFL levels measured using different platforms grouped by *APOE* genotypes in the BioFINDER-2 cohort. For each measurement, protein residuals eliminate the effect of age, sex, mean protein level (only in SomaLogic and OLINK) and Aβ status were used; significance was compared between each pair of genotypes, and FDR corrected. **G**. Heatmap shows the Spearman correlation between NEFL levels measured using different platforms in the BioFINDER-2 cohort. Each cell displays the Spearman correlation coefficient between two platforms, with both the color intensity and the number indicating the strength of the association (darker colors represent stronger correlations). Two-sided p-values were calculated and adjusted for multiple comparisons using the Benjamini–Hochberg (FDR) method. Statistically significant correlations after FDR correction are marked with an asterisk (*) next to the coefficient. **H**. The UpSet plot shows the intersection of proteins associated with *APOE4* in model 1 (without adjusting AD diagnosis or Aβ status) at each dataset using the unique gene symbol as Identifier. **I**. The

UpSet plot shows the intersection of proteins associated with *APOE2* in model 1 (without adjusting AD diagnosis or Aβ status) at each dataset using the unique gene symbol as Identifier.
