## Supplement Table 1 for "Proteomic signatures of the *APOE ε4* and *APOE ε2* genetic variants and Alzheimer’s disease"

| GNPC plasma SomaLogic (7285 proteins) | |
|---|---|
| | **N=4045** |
| **AD dementia diagnosis** | |
| CU | 2566 (63.4%) |
| AD | 1479 (36.6%) |
| **Sex** | |
| Male | 1667 (41.2%) |
| Female | 2378 (58.8%) |
| **Age** | |
| Mean (SD) | 73.3 (11.7) |
| Median [Min, Max] | 75.0 [20.0, 90.0] |
| ***APOE* genotypes** | |
| ε2ε2 | 13 (0.3%) |
| ε2ε3 | 375 (9.3%) |
| ε2ε4 | 111 (2.7%) |
| ε3ε3 | 1969 (48.7%) |
| ε3ε4 | 1334 (33.0%) |
| ε4ε4 | 243 (6.0%) |
| **Data contributor sites** | |
| C | 756 (18.7%) |
| F | 2233 (55.2%) |
| G | 129 (3.2%) |
| I | 52 (1.3%) |
| L | 528 (13.1%) |
| Q | 246 (6.1%) |
| R | 101 (2.5%) |

| BioFINDER-2 plasma SomaLogic (7285 protein | |
|---|---|
| | **N=1421** |
| **Aβ status (CSF Aβ42/40 ratio)** | |
| Aβ- | 715 (50.3%) |
| Aβ+ | 706 (49.7%) |
| **Sex** | |
| Male | 663 (46.7%) |
| Female | 758 (53.3%) |
| **Age** | |
| Mean (SD) | 67.8 (12.7) |
| Median [Min, Max] | 71.5 [20.0, 93.3] |
| ***APOE* genotypes** | |
| ε2ε2 | 5 (0.4%) |
| ε2ε3 | 90 (6.3%) |
| ε2ε4 | 44 (3.1%) |
| ε3ε3 | 524 (36.9%) |
| ε3ε4 | 629 (44.3%) |
| ε4ε4 | 129 (9.1%) |
| **AD dementia diagnosis** | |
| AD | 259 (18.2%) |
| CU | 846 (59.5%) |
| MCI | 316 (22.2%) |

| ADNI CSF SomaLogic (7001 proteins) | |
| --- | --- |
| | **N=666** |
| **Aβ status (Aβ-PET)** | |
| Aβ- | 280 (42.0%) |
| Aβ+ | 386 (58.0%) |
| **Sex** | |
| Male | 376 (56.5%) |
| Female | 290 (43.5%) |
| **Age** | |
| Mean (SD) | 73.3 (7.41) |
| Median [Min, Max] | 73.6 [54.4, 91.4] |
| ***APOE* genotypes** | |
| ε2ε2 | 1 (0.2%) |
| ε2ε3 | 45 (6.8%) |
| ~~ε2ε4~~ | ~~9~~ |
| ε3ε3 | 295 (44.3%) |
| ε3ε4 | 246 (36.9%) |
| ε4ε4 | 79 (11.9%) |
| **AD dementia diagnosis** | |
| AD | 123 (18.5%) |
| CN | 160 (24.0%) |
| MCI | 383 (57.5%) |

| BioFINDER-2 CSF OLINK (1391 proteins) | |
| --- | --- |
| | **N=1475** |
| **Aβ status (CSF Aβ42/40 ratio)** | |
| Aβ- | 699 (47.4%) |
| Aβ+ | 776 (52.6%) |
| **Sex** | |
| Male | 680 (46.1%) |
| Female | 795 (53.9%) |
| **Age** | |
| Mean (SD) | 68.4 (12.3) |
| Median [Min, Max] | 71.9 [20.0, 93.3] |
| ***APOE* genotypes** | |
| ε2ε2 | 4 (0.3%) |
| ε2ε3 | 93 (6.3%) |
| ε2ε4 | 45 (3.1%) |
| ε3ε3 | 533 (36.1%) |
| ε3ε4 | 667 (45.2%) |
| ε4ε4 | 133 (9.0%) |
| **AD dementia diagnosis** | |
| AD | 274 (18.6%) |
| CU | 846 (57.4%) |
| MCI | 355 (24.1%) |

| UKBB plasma OLINK (1319 proteins) | |
|---|---|
| | **N=4820** |
| **Sex** | |
| Male | 2599 (53.9%) |
| Female | 2221 (46.1%) |
| **Age** | |
| Mean (SD) | 54.2 (7.77) |
| Median [Min, Max] | 54.0 [40.0, 70.0] |
| ***APOE* genotypes** | |
| ε2ε2 | 29 (0.6%) |
| ε2ε3 | 592 (12.3%) |
| ε2ε4 | 106 (2.2%) |
| ε3ε3 | 2887 (59.9%) |
| ε3ε4 | 1104 (22.9%) |
| ε4ε4 | 102 (2.1%) |

| PPMI CSF OLINK (826 proteins) | |
|---|---|
| | **N=253** |
| **Sex** | |
| Male | 168 (66.4%) |
| Female | 85 (33.6%) |
| **Age** | |
| Mean (SD) | 63.5 (9.6) |
| Median [Min, Max] | 64.9 [30.6, 84.9] |
| ***APOE* genotypes** | |
| ε2ε2 | 2 (0.8%) |
| ε2ε3 | 28 (11.1%) |
| ~~ε2ε4~~ | ~~7~~ |
| ε3ε3 | 158 (62.5%) |
| ε3ε4 | 61 (24.1%) |
| ε4ε4 | 4 (1.6%) |
| **Diagnosis at baseline** | |
| PD | 72 (28.5%) |
| Prodromal | 56 (22.1%) |
| Healthy contol | 125 (49.4%) |
